# Leveraging electronic health records to examine differential clinical outcomes in people with Alzheimer’s Disease

**DOI:** 10.1101/2025.04.22.25326230

**Authors:** Shruthi Venkatesh, Linshanshan Wang, Michele Morris, Mohammed Moro, Ratnam Srivastava, Yunqing Han, Riddhi Patira, Sarah Berman, Oscar Lopez, Shyam Visweswaran, Tianrun Cai, Tianxi Cai, Zongqi Xia

## Abstract

**BACKGROUND:** Alzheimer’s disease (AD) carries a high societal burden inequitably distributed across demographic groups.

**OBJECTIVE:** To examine demographic differences and drivers of AD decline using real-world electronic health record (EHR) data with accurate AD population identification.

**METHODS:** Leveraging EHR data (1994-2022) from two large independent healthcare systems, we applied a novel unsupervised phenotyping algorithm to predict AD diagnosis and validated using gold-standard chart-reviewed and registry-derived diagnosis labels. Among patients with ≥24 months of EHR data not living in nursing homes pre-AD diagnosis, we performed healthcare system-specific competing risk survival analyses to estimate the time to two readily ascertainable AD decline outcomes (*i.e.,* nursing home admission, death), stratified by demographic groups and accounting for baseline covariates (*e.g.,* age, gender, race, ethnicity, healthcare utilization, and comorbidities). We then performed a covariate-adjusted fixed-effects meta-analysis using inverse variance weighting to estimate the time to AD decline stratified by demographic groups.

**RESULTS:** The algorithm achieved robust prediction in identifying AD patients across both healthcare systems (AUROC score range: 0.835-0.923) and demographic groups. Of the 29,262 AD patients in both healthcare systems (61% women, 90% non-Hispanic White, 79.52±9.39 years of age at AD diagnosis), 49% entered nursing homes and 48% died during follow-up. In covariate-adjusted fixed-effects meta-analysis, women had a higher risk of nursing home admission (HR[95% CI]=1.061[1.024-1.100], *p*=.001) and lower risk of death (HR[95% CI]=0.856[0.811-0.904], *p*<.0001) than men. Non-Hispanic White individuals had similar nursing home risk (HR[95% CI]=1.006[0.952-1.063], *p*=0.831) but higher death risk (HR[95% CI]=1.376[1.245-1.521], *p*<.0001) than racial and ethnic minorities. Older age at AD diagnosis and greater pre-existing comorbidity burden increased both nursing home admission and death risk.

**CONCLUSION:** Findings from two large EHR cohorts add to the real-world evidence of demographic differences in clinical AD decline and its drivers, which could potentially inform individual clinical management and future public health policies.

## INTRODUCTION

Late-onset Alzheimer’s disease (AD) is the leading cause of dementia and neurological disability affecting ∼7.2 million people in the United States.^1–3^ People with AD experience variable rates of decline with differences among demographic groups. The likelihood of nursing home admission, indicating decline in capacity for independent living, is higher among women and racial and ethnic minorities with AD.^4–7^ In contrast, post-diagnosis survival is shorter for men and non-Hispanic White (NHW) individuals.^5,8–10^

Prior epidemiological or claims-based studies implicated sociodemographic factors and comorbidities as potential drivers of differential AD decline.^11–16^ Comprehensive assessment of these modifiable factors and quantifying the extent to which they drive AD decline using large-scale *real-world* clinical data can augment prior research and enhance generalizability of the findings. Specifically, electronic health record (EHR) data containing the longitudinal clinical profiles of demographically broad patient populations could complement traditional data sources (*e.g.,* epidemiological studies that are costly and time-consuming to conduct, claims data that lack clinical granularity) given the growing adoption and standardization of EHR. Notably, EHR data have advantages over claims data (*e.g.,* Medicare) to examine differential AD decline. First, unstructured narrative data (*e.g.,* clinical narratives that contextualize diagnosis, treatment, and prognosis) adds significant value beyond structured codified data (*e.g.,* diagnoses and prescriptions). Second, EHR data can be more readily linked to local registry data (*e.g.,* Alzheimer’s Disease Research Center) that provide the critical gold-standard labels for developing algorithms applicable at the point of care and for scaling to examine non-registry patients in the EHR. Third, EHR data can undergo the crucial local quality control (*e.g.,* chart review), which would typically be infeasible for claims data.

Despite the advantages of EHR, accurate identification of target populations (*e.g.,* AD) from the EHR has limited the effective use of EHR data for examining drivers of differential AD decline (**S-Introduction-1**). While rule-based approaches (*e.g.,* identifying cases using diagnosis codes, diagnostic tests, prescriptions or the combination thereof) have achieved reasonable performance in identifying AD populations in claims data, gold-standard ground truth is rarely available with rare exceptions.^17–21^ On the other hand, heterogeneity in diagnosis coding and clinical documentation leads to suboptimal performance of rule-based approaches in accurately identifying AD populations in the EHR.^22–25^ Limitations of existing approaches to identify AD populations from the EHR include sparse gold-standard labels to validate AD diagnosis status (particularly via external validation) and the lack of validation across demographic groups despite known differences in clinical presentation.^25–36^

Here, we re-examined the demographic differences and drivers of AD decline using real-world EHR data with accurate AD population identification. We leveraged the EHR data of two independent healthcare systems, one of which was linked to an AD registry, applied a knowledge graph-guided unsupervised phenotyping algorithm to predict AD diagnosis across demographic groups, and validated algorithm performance using chart-reviewed and registry-derived gold-standard labels. Using these large real-world AD-EHR cohorts, we then examined differences in two *readily ascertainable* clinical outcomes of AD decline (*i.e.,* nursing home admission, mortality) between demographic groups (*e.g.,* race and ethnicity, gender) and explored potential drivers of differential AD decline.

## METHODS

### Ethics Approval

The Institutional Review Board of the University of Pittsburgh (STUDY21020153) and Mass General Brigham (MGB, 2023P001450) approved the study protocol. The study using de-identified data was deemed exempt.

### Harmonizing EHR data and creating AD-EHR data-marts

We obtained inpatient and outpatient EHR data from two healthcare systems, UPMC (Pittsburgh, PA; codified data: 2004-2022; narrative data: 2011-2022) and MGB (Boston, MA; codified and narrative data: 1994-2022). Codified data contained demographics, diagnoses (*e.g.*, International Classification of Disease [ICD] code), procedures (*e.g.*, Current Procedural Terminology [CPT] code), healthcare utilization metrics (*e.g.*, total number of ICD codes and encounter notes), medication prescriptions (*e.g.*, RxNorm codes), and laboratory tests (*e.g.*, Logical Observation Identifiers Names and Codes [LOINC] codes).^37,38^ We organized diagnoses by mapping ICD-9 and ICD-10 codes to PheCodes,^39,40^ consolidated procedures using the Clinical Classifications Software for Services and Procedures (Agency for Healthcare Research and Quality), and mapped electronic prescriptions to RxNorm ingredient level. We deployed an established natural language processing pipeline (Narrative Information Linear Extraction [NILE]) to derive concept unique identifiers (CUIs) from clinical narratives (*e.g.,* physician office notes, discharge summaries) according to unified medical language system (UMLS).^41,42^ Aiming high sensitivity to start, we built the *initial* **AD-EHR data-marts**, comprising UPMC (2011- 2022) and MGB (1994-2022) patients with ≥1 ICD code for AD or related dementia (*e.g.,* ICD-9=290.x, 294.2x, 331.0; ICD-10=F03.9x, G30.x; **S-Table 1, S-Method-1**). We imputed missing race and ethnicity using age and gender.^43,44^ We performed sensitivity analyses by excluding patients from the initial AD-EHR data-marts with missing race and ethnicity information (**S-Method-2, S-Figure 1**).

**Figure 1.**
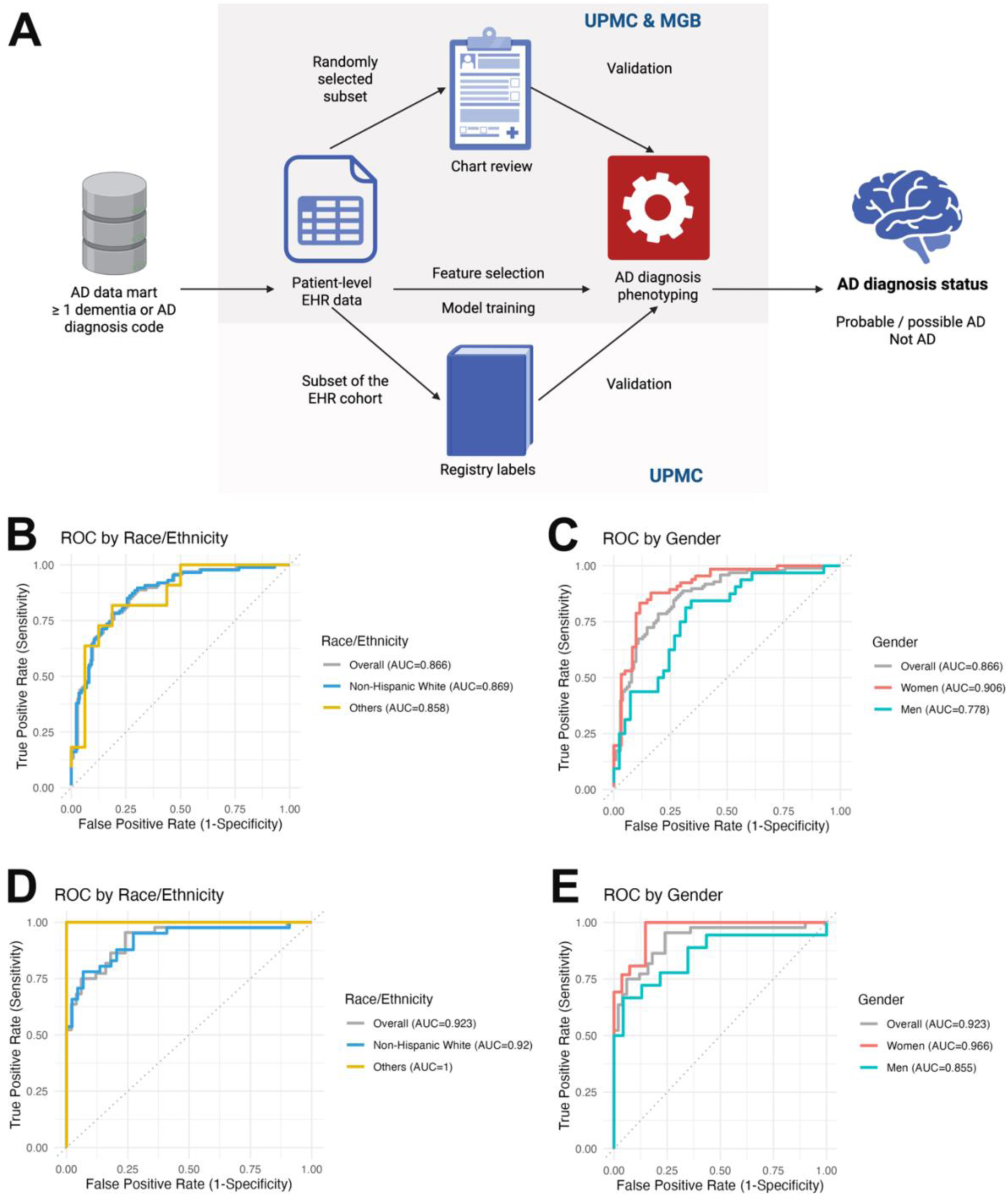
AD diagnosis phenotyping algorithm. (A) Schematic overview. (B-C) Phenotyping algorithm performance at UPMC. (D-E) Phenotyping algorithm performance at MGB.

**Table 1.**
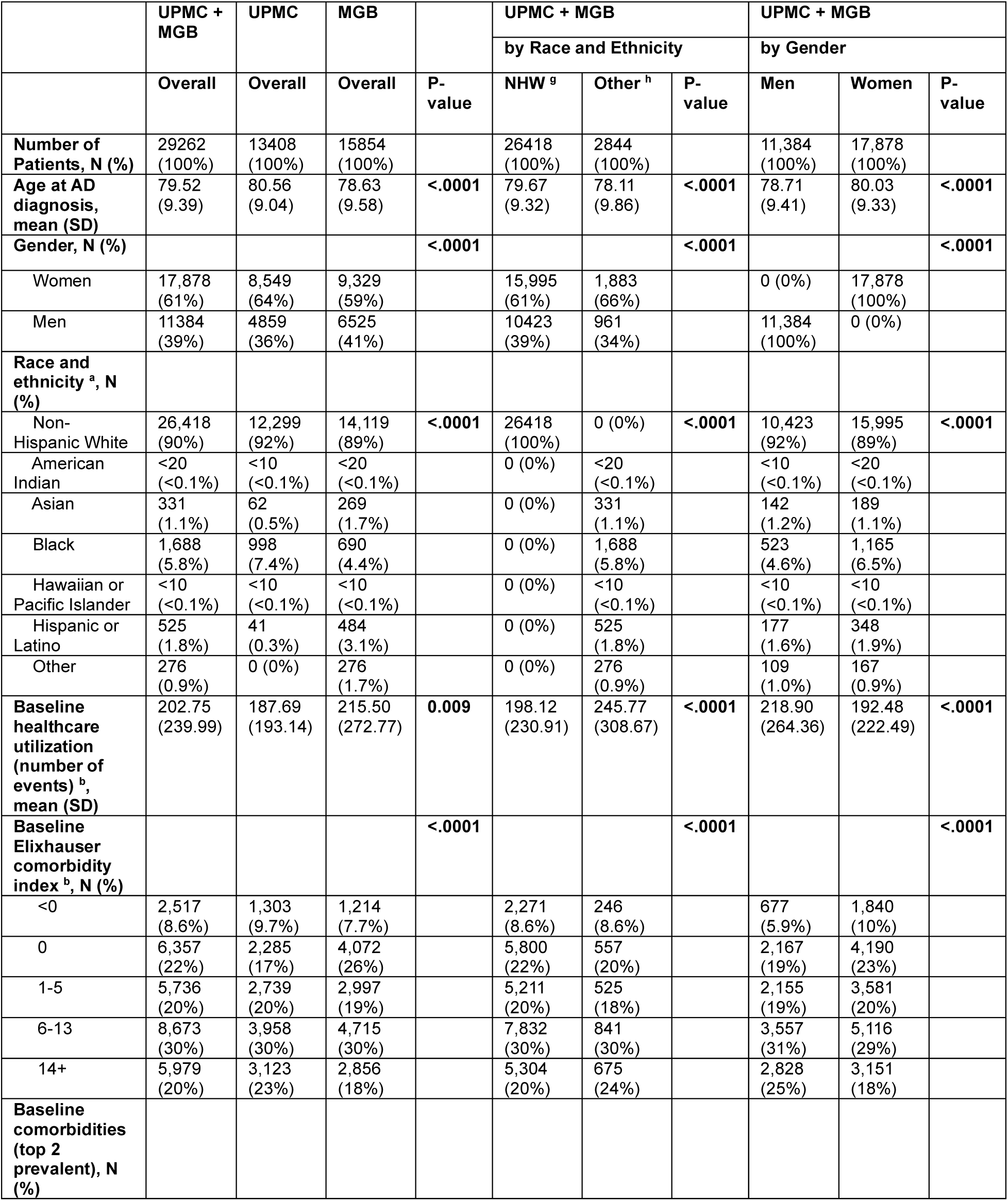
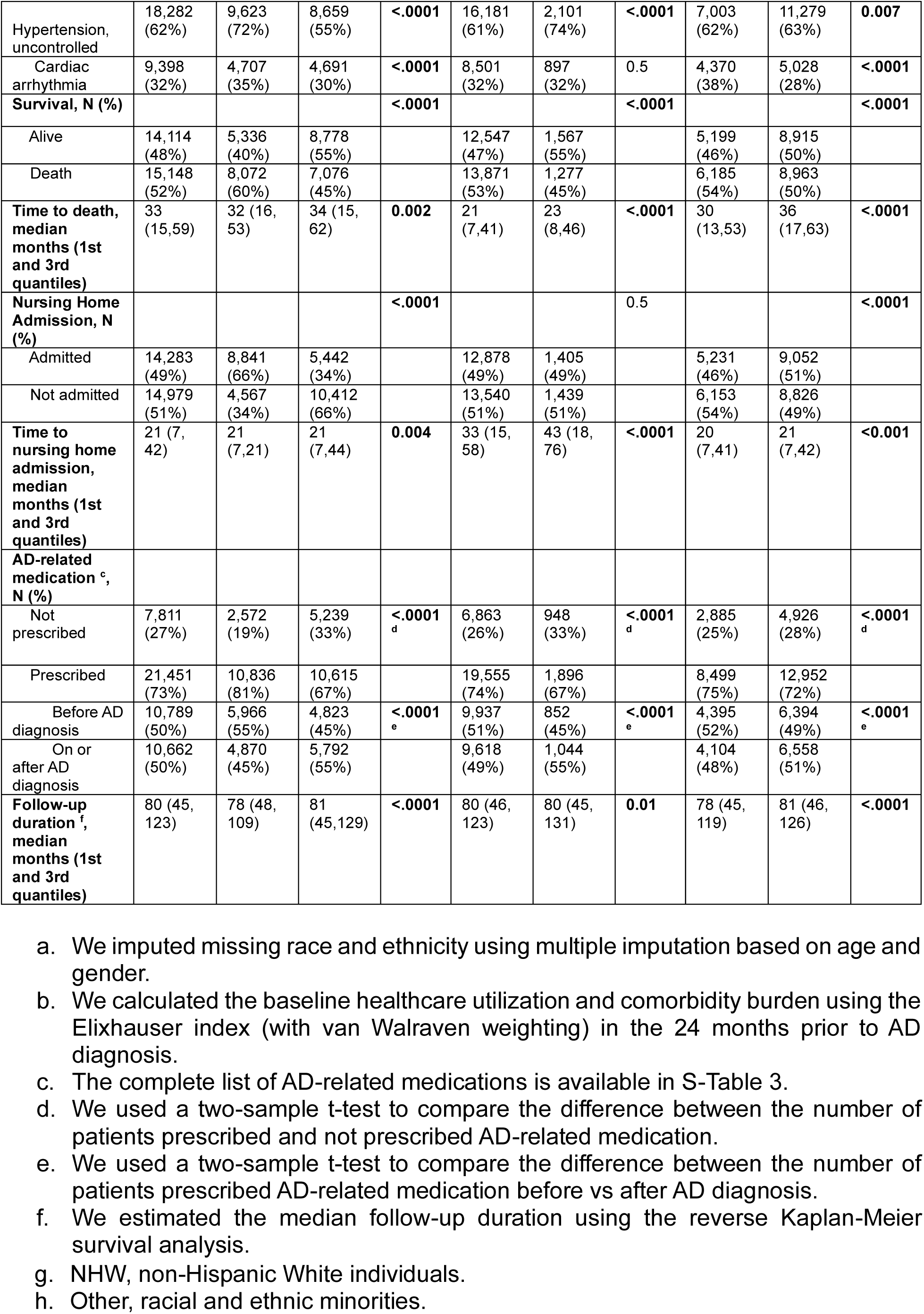
Cohort characteristics.

### Inclusion Criteria

We applied a novel knowledge graph guided unsupervised phenotyping algorithm, <u>K</u>nowledge driven <u>O</u>nline <u>M</u>ultimodal <u>A</u>utomated <u>P</u>henotyping (**KOMAP**), to assign AD diagnosis (*i.e.,* probable or possible AD vs *not* AD) for all patients in the initial AD-EHR data marts (**Figure 1A, S-Table 2**).^45^ We validated KOMAP-predicted AD diagnosis status using gold-standard chart-reviewed labels (UPMC, n=200; MGB, n=100, **S-Method-2**) and registry labels (University of Pittsburgh Alzheimer’s Disease Research Registry [ADRC], n=1916 with UPMC EHR linkage) across demographic groups. We classified individuals from AD-EHR data-marts with KOMAP-predicted AD diagnosis at 90% specificity as having AD (**Figure 2A**, **S-Method-2**). We held the specificity at 90% to minimize false positives while keeping reasonably large sample sizes. The first AD PheCode (*i.e.,* PheCode 290.11) occurrence was the index date or *baseline*. For downstream analyses, we excluded patients with <24 months of EHR data or admitted to nursing homes (*i.e.,* with ≥1 diagnosis code ***or*** CUI for nursing home admission) prior to the baseline.

**Figure 2.**
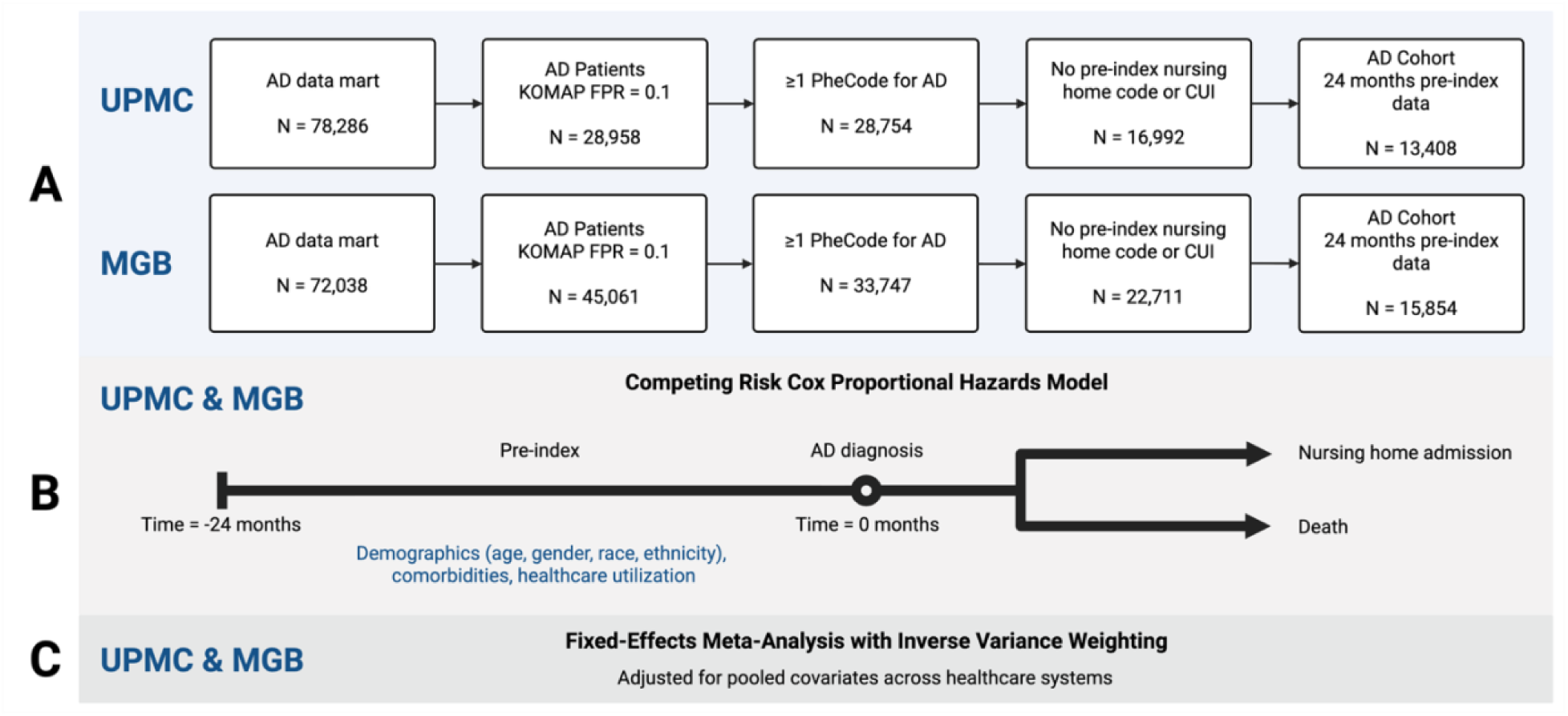
Study design. (A) Inclusion criteria. (B) Healthcare system-specific covariate-adjusted Cox proportional hazard models. (C) Fixed-effects meta-analysis of healthcare system-specific covariate-adjusted Cox proportional hazard models.

### Covariates

We accounted for covariates, including demographics (*e.g.*, age at AD diagnosis, gender, race, ethnicity) and *baseline* clinical profiles (*e.g.,* healthcare utilization, pre-existing comorbidity burden) in the 24 months preceding the index date. Age at AD diagnosis was operationally defined as the age at the first AD PheCode. Self-reported gender included men and women. Race included American Indian or Alaska Native, Asian, Black, Native Hawaiian or Pacific Islander, White, and others. Ethnicity included Hispanic or Latino and non-Hispanic. We dichotomized race and ethnicity as non-Hispanic White (*i.e.,* NHW) versus other minorities, which included individuals of Hispanic and non-European descent. Baseline healthcare utilization was the annualized total number of diagnosis codes and clinical encounters.^46^ Baseline comorbidity burden was the Elixhauser comorbidity index (ECI).^47^ Using the R *comorbidity* package, we mapped diagnosis codes to the 29 pre-existing health conditions comprising the ECI.^48^ We considered each comorbidity as an individual covariate in the main analysis, and performed a sensitivity analysis using the ECI score (**S-Method-2**).^49^ We included the most common and consistently available covariates in the EHR (*e.g.,* demographics, comorbidities, healthcare utilization) relevant to AD to balance clinical relevance and data completeness. While incorporating other factors (*e.g.,* social determinants of health, blood or neuroimaging biomarkers) may reduce residual confounding, they are currently not readily ascertainable from the EHR for most patients.

### Clinical outcomes of AD decline

We assessed two clinically relevant indicators of AD decline *readily ascertainable* from EHR: (1) time to nursing home admission and (2) time to death. Nursing home admission is routinely documented in clinical practice.^6,7^ We defined nursing home admission as having ≥1 code *or* CUI for admission to or clinical encounters in any type of residential institution (*e.g.,* skilled nursing facility, long-term care facility, end-of-life care facility, **S- Table 3**), validated with gold-standard chart-reviewed labels (**S-Method-3**). Time to nursing home admission was from the index date to the first occurrence of any nursing home code or mention. Death status in EHR was linked to the social security death index. Time to death was from the index date to death. For patients without reaching either endpoint, we used all available EHR data.

### Statistical analysis

We used two-tailed t-tests or two-proportion Z-tests to compare differences in demographics and baseline clinical profiles (**S-Table 4**) and prevalence of AD clinical outcomes among demographic groups (**S-Method-4**). We estimated the time-to-outcome using healthcare system-specific covariate-adjusted Cox proportional hazards (PH) models that accounted for the competing risks of nursing home admission and death, stratified by demographic groups (**Figure 2B**). Using patient-level data from both sites, we performed a fixed-effects meta-analysis using inverse variance weighting to estimate the time-to-outcome stratified by demographic groups, adjusting for pooled covariates (**Figure 2C)**. (Random-effects models would be unstable with two sites).

### Data and code availability

Codes for KOMAP and project analysis are available on Github (**S-Method-5**).^45,50^

## RESULTS

### AD diagnosis phenotyping algorithm performance

Using chart-reviewed labels (**S-Table 5**) to evaluate AD diagnosis phenotyping algorithm performance, KOMAP achieved a higher AUROC at MGB (n=100, 45% AD, AUROC=0.923) than UPMC (n=200, 49% AD, AUROC=0.854, **Figure 1B-E, S-Tables 6-9, S-Result-1**). While KOMAP achieved higher AUROC in women at both healthcare systems (UPMC: women=0.888, men=0.782; MGB: women=0.965, men=0.855), there was inconsistent pattern between two systems for race and ethnicity groups (UPMC: NHW=0.856, other=0.854; MGB: NHW=0.925, other=1.000). Using registry-derived labels to evaluate algorithm performance at UPMC (n=1916, 47% AD, AUROC=0.835, **S- Table 10**), KOMAP achieved higher AUROC in women than men and higher in racial and ethnic minorities than NHW individuals (women=0.846, men=0.824; NHW=0.821, other=0.922). In sensitivity analysis (without imputation of missing race and ethnicity), the algorithm achieved similar performances (**S-Table 11**).

### AD cohort characteristics

The combined AD cohort comprised 29,262 patients (61% women, 90% NHW, mean[SD]=79.52[9.39] years at AD diagnosis, **Table 1**), including 13,408 UPMC patients (64% women, 92% NHW, 80.56[9.04] years at AD diagnosis) and 15,854 MGB patients (59% women, 89% NHW, 78.63[9.58] years at AD diagnosis). AD cohorts in the two healthcare systems differed in characteristics. Compared to MGB patients, UPMC patients had older age at AD diagnosis, greater proportion of women and NHW individuals, lower baseline healthcare utilization, greater pre-existing comorbidity burden, greater proportion of nursing home admission and death during study follow-up, greater proportion of AD-related medication prescriptions (particularly before AD diagnosis), and shorter follow-up duration (**Table 1, S-Table 12, S-Result-2**).

In the combined AD cohort, demographic groups differed in patient profiles (**Table 1**). At AD diagnosis, women were older than men, while NHW individuals were older than racial and ethnic minorities (mean[SD] years: women=80.03[9.33], men=78.71[9.41], *p*<.0001; NHW=79.67[9.32], other=78.11[9.86], *p*<.0001). Baseline healthcare utilization was higher in men than women and higher in racial and ethnic minorities than NHW individuals (mean[SD] healthcare utilization counts: women=192.48[222.49], men=218.90[264.36], *p*<.0001; NHW=198.12[230.91], other=245.77[308.67], *p*<.0001). The pre-existing comorbidity burden was overall high, with men having higher burden than women and racial and ethnic minorities having greater burden than NHW individuals, respectively (**Table 1, S-Figure 2**). While most patients were prescribed AD-related medications, women and racial and ethnic minorities were prescribed at lower rates than men and NHW individuals, respectively (n[%] AD-related prescriptions: overall=21,451[73%]; women=12,952[72%], men=8,499[75%], *p*<.0001; NHW=19,555[74%], other=1,896[67%], *p*<.0001, **Table 1**). During the study follow-up period, 14,283 (49%) patients were admitted to nursing homes and 15,148 (52%) died. While a higher proportion of women than men were admitted to nursing homes, a higher proportion of men and NHW individuals died during follow-up than women and racial and ethnic minorities (n[%] admitted to nursing homes: women=9,052[51%], men=5,231[46%], *p*<.0001; NHW=12,878[49%], other=1,405[49%], *p*=.05; n[%] death: women=8,963[50%], men=6,185[54%], *p*<.0001; NHW=13,871[53%], other=1,277[45%], *p*<.0001). In sensitivity analysis (without imputation of missing race and ethnicity), patient profiles were comparable to the main AD cohort (**S-Table 13, S-Result-3**).

### Differential risk of nursing home admission and death

Using fixed-effects meta-analysis of the healthcare system-specific covariate-adjusted Cox-PH models, we compared the risk of two pragmatic clinical outcomes among demographic groups. First, women had a higher risk of nursing home admission than men (HR [95% CI]=1.061 [1.024-1.100], *p*=.001, **Figure 3, S-Table 14, S-Result-4**), while there was no significant differences between race and ethnicity groups (HR [95% CI]=1.006 [0.952-1.063], *p*=0.831). Second, NHW individuals had a higher risk of death than racial and ethnic minorities (HR [95% CI]=1.376 [1.245-1.521], *p*<.0001, **S-Table 15**), while women had a lower risk than men (HR [95% CI]=0.856 [0.811-0.904], *p*<.0001).

**Figure 3.**
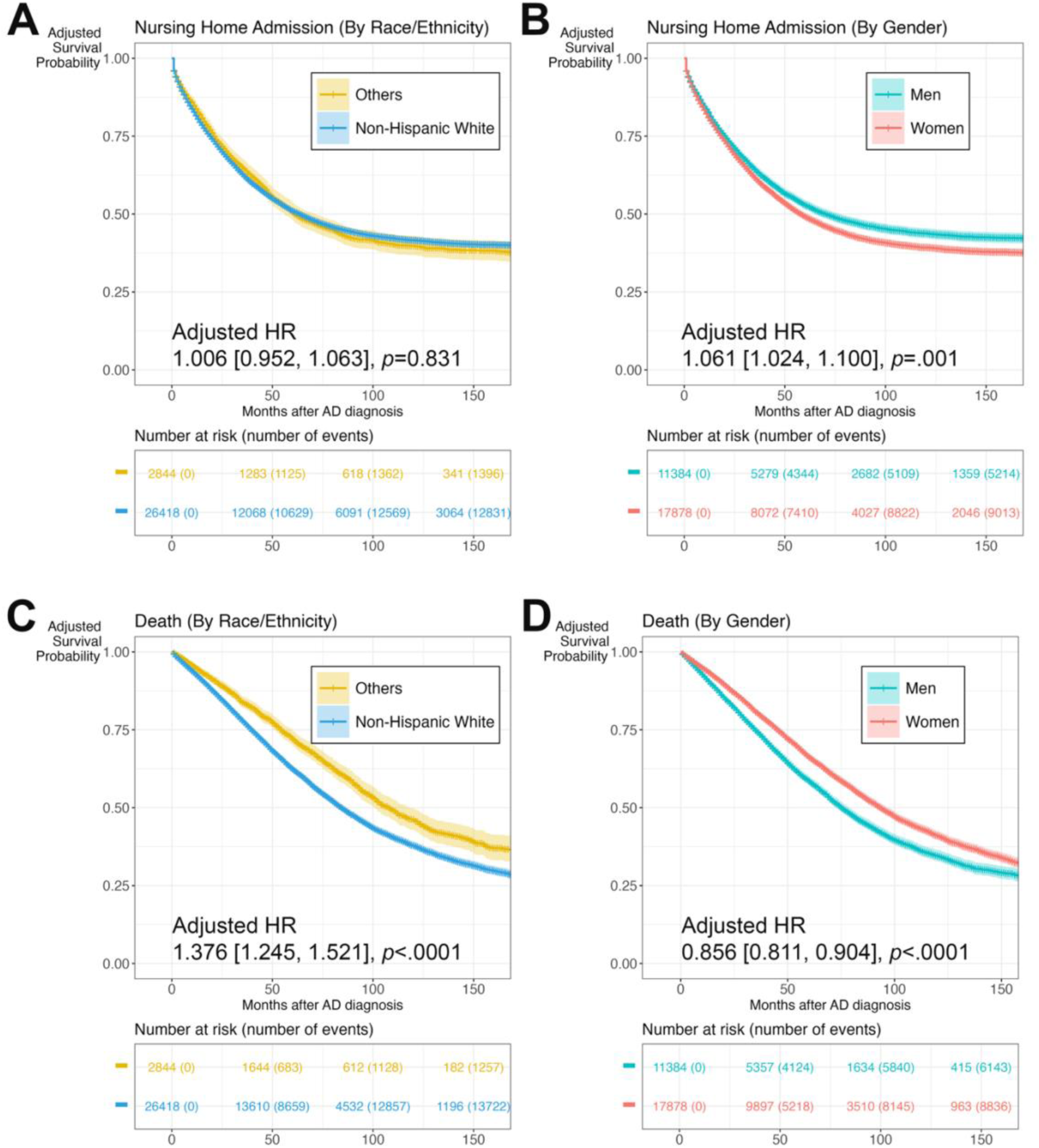
Fixed-effects meta-analysis of healthcare system-specific covariate-adjusted Cox proportional hazard models to estimate time to nursing home admission and death by demographic groups. The fixed-effects meta-analysis model was adjusted for pooled covariates across both healthcare systems from inverse-probability weighting. Covariates included demographics (*e.g.*, age at AD diagnosis, gender, race, ethnicity) and baseline clinical profiles (*e.g.,* healthcare utilization, pre-existing comorbidity burden) in the 24 months preceding the index date. (A-B) Nursing Home Admission. (C-D) Death. Reference group for race and ethnicity: non-Hispanic White. Reference group for gender: women.

Key drivers of the increased nursing home admission risk (**Figure 4A**) included older age at AD diagnosis, being a woman, high baseline healthcare utilization, and high pre-existing comorbidity burden characterized by the presence of psychiatric disorders (*e.g.,* psychoses, depression), cardiometabolic and pulmonary disorders (*e.g.,* hypertension, diabetes, chronic pulmonary disease), or renal disorders (*e.g.,* fluid and electrolyte disorders). Key drivers of the increased death risk (**Figure 4B**) included older age at AD diagnosis, being male, being NHW, low healthcare utilization, and high pre-existing comorbidity burden characterized by the presence of neoplastic disorders (*e.g.,* metastatic cancer, solid tumor without metastasis), cardiometabolic and vascular disorders (*e.g.,* diabetes, congestive heart failure, peripheral vascular disease), or psychiatric disorders (*e.g.,* psychoses). Sensitivity analyses using a single score for pre-existing comorbidity burden and excluding patients with missing race and ethnicity information yielded similar results (**S-Figures 3-6**, **S-Table 14**, **S-Table 15**).

**Figure 4.**
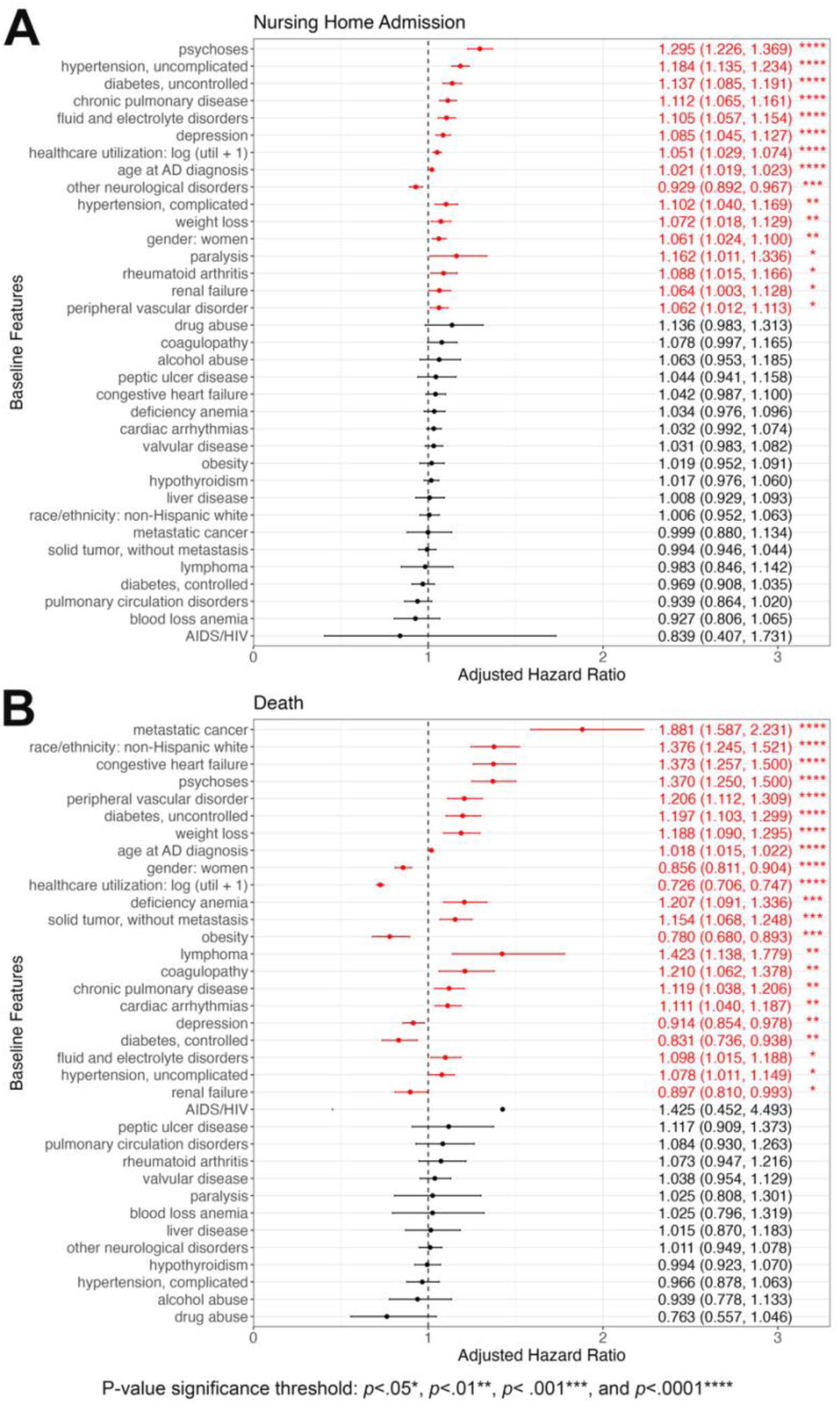
Adjusted hazard ratios of variables in the fixed-effects meta-analysis of healthcare system-specific covariate-adjusted Cox proportional hazard models of nursing home admission and death. The fixed-effects meta-analysis model was adjusted for pooled covariates across both healthcare systems from inverse-probability weighting. Covariates included demographics (*e.g.*, age at AD diagnosis, gender, race, ethnicity) and baseline clinical profiles (*e.g.,* healthcare utilization, pre-existing comorbidity burden) in the 24 months preceding the index date. Variables are ordered first by p-value significance threshold (*p*<.05*, *p*<.01**, *p*<.001***, *p*<.0001****) and then effect size. (A) Nursing Home Admission. (B) Death.

## DISCUSSION

We re-examined the previously reported demographic differences and drivers of AD decline using real-world clinical data from two large healthcare systems with accurate AD population identification. We identified AD populations using a novel unsupervised phenotyping algorithm (KOMAP), which achieved *overall* robust predictive performance with modest differences across healthcare systems and demographic groups (**S- Discussion-1**). Demographic groups in the AD-EHR cohorts exhibited differential risks of clinical decline. Women had a higher risk of nursing home admission, while NHW individuals and men had a higher risk of death. Older age at AD diagnosis and greater pre-existing comorbidity burden increased both nursing home admission and death risk.

In these real-world AD-EHR cohorts, the risk of AD decline differed across demographic groups, largely consistent with prior epidemiological or claims-based studies.^4–10^ Women had a higher risk of nursing home admission than men, possibly due to longer life expectancy.^4,5,35,51^ In contrast, men had a higher risk of death than women, potentially attributable to the greater pre-existing comorbidity burden in men, which would be consistent with their higher baseline healthcare utilization. Further, men with AD might have a greater AD symptom burden (not measured here), leading to more rapid decline.^52^ While prior studies (using claims data) found racial and ethnic minorities having a higher risk of nursing home admission than NHW individuals, our study did not confirm this finding.^6,7^ Interestingly, NHW patients had a higher risk of death than racial and ethnic minorities, mirroring the broader pattern in “mortality crossover” among older adults.^8–10,16,53^ As a possible explanation, racial and ethnic minorities more likely remained undiagnosed, underestimating their AD-related mortality.^54^ This study also cannot address whether biological factors drive differences in clinical outcomes across demographic groups.^55^

We found older age at AD diagnosis and greater pre-existing comorbidity burden (particularly psychiatric and cardiometabolic disorders) as key drivers of higher risks of *both* nursing home admission and death.^11–16,56^ Given that healthcare utilization and comorbidities generally increase with age, older age of diagnosis may reflect diagnostic delay (rather than later disease onset), possibly explaining its role in worse AD outcomes. Greater pre-existing comorbidity burden compounds AD management challenges and increases comprehensive and specialized care needs, which are inaccessible to many patients. Interestingly, high baseline healthcare utilization (likely reflecting greater baseline functional impairment) increased nursing home admission risk, whereas low healthcare utilization (possibly reflecting inadequate symptom and comorbidity management) increased death risk. Taken together, clinical investigations using real-world AD cohorts would benefit from using multiple interrelated endpoints and careful interpretations.

The study has several strengths. First, we overcame the limitations of existing approaches to identifying AD populations (*e.g.,* sparse gold-standard labels, lack of validation across demographic groups, lack of external validation) by validating the algorithm with gold-standard labels in two independent healthcare systems and across demographic groups (**S-Discussion-2**).^25–34^ The overall robust performance in identifying people with AD supports our AD cohorts as a valuable clinical research resource. Second, we demonstrated the feasibility of using these two large AD-EHR cohorts with long-term follow-up to examine the time to two pragmatic but clinically relevant endpoints, *i.e.,* nursing home admission and death. These real-world AD cohorts derived data from both academic and community practices within two geographically distinct large catchment areas. Moreover, these real-world AD cohorts are more representative of clinical AD populations than traditional epidemiological studies. Third, although >40% of people with AD eventually reside in nursing homes, nursing home admission has been underutilized in EHR-based AD studies.^26,57–59^ We applied a rule-based definition (≥1 code or CUI related to nursing home) to identify nursing home admission from the EHR, which was validated using gold-standard chart-reviewed labels. While other key clinical outcomes such as cognition and function fluctuate over time, nursing home admission and death are concrete milestones readily ascertainable from the EHR. Finally, we comprehensively assessed key demographic factors and pre-existing comorbidities and quantified the extent to which drive AD decline. Specifically, we assessed common comorbidities using ECI, a well-validated measure of comorbidity burden.

Our study also has limitations. First, we grouped multiple racial and ethnic groups (*e.g.,* American Indian, Asian, Black, Hispanic) as one minority group due to the modest sample size of each. While statistically necessary, this approach might obscure meaningful group differences and limit the generalizability to these demographic groups. Second, in this first series of studies using these AD-EHR cohorts, we did not assess cognitive or functional decline. Given the sparse EHR documentation of cognition (*e.g.,* Mini Mental Status Exam, Montreal Cognitive Assessment) and function (*e.g.,* Functional Activities Questionnaire) status,^60^ we are developing novel methods to impute these longitudinal measures for future studies. Third, these AD cohorts may include probable and possible AD and various clinical presentations (*e.g.,* amnestic, nonamnestic). Additional methods will be necessary to subtype the heterogeneous AD population. Through linkage with ADRC (at UPMC), we will compare the algorithm-predicted AD diagnosis with biomarker-informed AD diagnosis in the future. Finally, biases inherent in EHR data such as potential data incompleteness (*e.g.,* leakage due to the fragmented healthcare landscape) and selection bias (*e.g.,* stemming from disparities in diagnosis rates, healthcare access, and survival) might influence results of AD diagnosis phenotyping and estimation of AD decline.^61^

## CONCLUSION

We created two large real-world AD cohorts with long follow-up in two independent healthcare systems. This cohort study underscored the demographic differences in the risk of nursing home admission and death among the AD population and identified potential drivers. Future research using such real-world cohorts will examine the contribution of additional factors (*e.g.,* social determinants of health as their coding are gradually introduced into routine clinical care) beyond demographics, healthcare utilization and comorbidities, deploy formal causal inference approaches, and incorporate imputed cognitive, functional and frailty status to complement nursing home admission and death as outcomes. This research can inform targeted interventions to modify the risk of AD decline.

## Data Availability

Codes for KOMAP and project analysis are available on Github. We will publicly disseminate anonymous summary-level registry data and EHR data. The rationale for not sharing patient-level data is that patient-level clinical data (either de-identified information or limited protected health information containing dates of clinical events or even if anonymous due to concern for re-identification) are universally subject to the rules and regulation of each healthcare system, which may only be affiliated with but are not the same as the primary academic institutions of the study investigators. Sharing of de-identified EHR data with qualified external researchers by each of the study performance site may be permissible only after the approval of the respective Institutional Review Boards (IRBs), regulatory oversight agents of the healthcare systems (that own the clinical data) as well as the appropriate Data Usage Agreements (DUA) between institutions.

https://github.com/xialab2016/adphenotyping

https://github.com/xinxiong0238/KOMAP

https://shiny.parse-health.org/ONCE/

## ACKNOWLEDGEMENTS

We would like to thank the patients whose data contributed to the research findings.

This study was supported by the National Institutes of Health under award numbers R01 NS098023 and R01 NS124882 from the National Institute of General Medical Sciences. The content is solely the responsibility of the authors and does not necessarily represent the official views of the National Institutes of Health.

