## Supplementary Material for "Leveraging electronic health records to examine differential clinical outcomes in people with Alzheimer’s Disease"

### SUPPLEMENTARY INTRODUCTION

**S-Introduction-1. Motivation for identifying AD populations using a novel AD diagnosis phenotyping algorithm**

Existing approaches to identify AD populations from EHR have limitations. Using diagnosis codes alone yields poor accuracy due to lack of specificity for AD compounded by diagnostic delay.[1–3] Recent efforts using structured codified data and/or unstructured clinical information (*i.e.,* clinical narrative) largely relied on the EHR data of a single healthcare system and lacked external validation.[4–12] To validate the AD diagnosis phenotyping algorithms, prior studies used labels such as clinical assessment, chart review, and surrogate indicators within the EHR.[4] Few AD-EHR cohorts hold the crucial linkage to AD research registries that contain gold-standard diagnosis labels for validating the algorithm performance. Finally, to our knowledge, no prior AD diagnosis phenotyping efforts validated performance across demographic groups despite the known differences in clinical presentations, which may exacerbate algorithm bias. Taken together, this leaves an unmet need for more accurate identification of AD populations from the EHR with robust validation of algorithm performance.

### SUPPLEMENTARY METHODS

**S-Method-1. Harmonizing EHR data and creating AD-EHR data-marts**

From the codified and narrative EHR data, we built the *initial* AD-EHR data-marts, comprising UPMC and MGB patients with ≥1 ICD code for AD or related dementia (*e.g.,* ICD-9=290.x, 294.2x, 331.0; ICD-10=F03.9x, G30.x; **S-Table 1**). As narrative data were collected at UPMC only after January 1, 2011, we included UPMC patients with codified and narrative data from 2011 to 2022. We included MGB patients with codified and narrative data from 1994 to 2022.

**S-Method-2. Identifying AD diagnosis and creating AD cohorts**

Using an unsupervised phenotyping algorithm, Knowledge driven Online Multimodal Automated Phenotyping (**KOMAP**),[13] we assigned AD diagnosis (*i.e.,* probable or possible AD vs *not* AD) for all patients in the initial AD-EHR data marts (**main Figure 1A**). First, KOMAP leveraged an online narrative and codified feature search engine (ONCE) powered by multi-source knowledge-graph representation learning to generate a list of informative codified and narrative features relevant to the target clinical concept, *i.e.,* AD (**S-Table 2**). Second, using the same list of ONCE-selected features, we trained KOMAP independently at UPMC and MGB to account for population heterogeneity. We combined probable and possible AD into a single category for prediction of AD diagnosis status as they represented the heterogeneous presentations of AD. Although KOMAP itself is unsupervised, we evaluated the performance of the AD phenotyping algorithm in predicting AD diagnosis status using gold-standard labels. To validate KOMAP-predicted AD diagnosis, we obtained gold-standard chart-reviewed labels (UPMC, n=200; MGB, n=100) in addition to registry-derived labels (University of Pittsburgh Alzheimer's Disease Research Registry [ADRC], n=1916 with UPMC EHR linkage) for a subset of the patients in the initial AD-EHR data marts (comprising patients ≥1 dementia or AD PheCode).

*S-Method-2.1. Validation of KOMAP-predicted AD diagnosis status with chart-reviewed labels*

After excluding AD patients enrolled in the University of Pittsburgh Alzheimer's Disease Research Center (ADRC) registry, we randomly sampled 200 patients from the UPMC AD data mart for chart review. We also randomly sampled 100 patients from the MGB AD data mart for chart review. We sampled fewer patients from MGB due to better clinical documentation. After training by an experienced cognitive and behavioral neurologist (RP), a clinical domain expert (SV) reviewed the EHR of these patients to determine AD diagnosis status and used the annotated labels as the gold-standard ground truth for validating KOMAP-predicted AD diagnosis status. We applied the 2011 diagnostic criteria for AD in the chart review.[14] Patients whose clinical documentation met the core clinical criteria for probable or possible AD were classified as having a diagnosis of AD. Patients who did not meet these criteria were classified as not having AD. We did not assess any measures of chart review reliability or validity since one clinical domain expert reviewed the medical records.

*S-Method-2.2. Validation of KOMAP-predicted AD diagnosis status with registry-derived labels*

The University of Pittsburgh ADRC has maintained a longitudinal prospective registry of AD patients for over 25 years. For the 1,916 patients in the UPMC AD data mart who were enrolled in the ADRC registry, we linked their EHR data with ADRC registry elements (including clinician-determined AD diagnosis status) using an honest broker system. We validated KOMAP-predicted AD diagnosis status with clinician-determined AD diagnosis status from the ADRC registry.

*S-Method-2.3. Comparison of KOMAP performance to benchmark phenotyping methods*

We also compared the performance of KOMAP to common benchmark phenotyping methods constructed from surrogate features predictive of AD diagnosis. These surrogate features included the number of PheCodes for "Alzheimer's disease" (PheCode.290.11), "dementia" (PheCode.290.1), "delirium, dementia, and amnestic and other cognitive disorders" (PheCode.290), and the number of mentions of the main CUI (*i.e.,* narrative feature) for "Alzheimer's disease" (C0002395). Further, we trained KOMAP separately by race and ethnicity and by gender groups ("KOMAP separate"). We compared the performance of KOMAP and KOMAP separate to assess if phenotyping needed to be performed independently based on demographic factors.

*S-Method-2.4. Phenotyping algorithm performance*

We obtained performance measures of all phenotyping methods stratified by race and ethnicity and by gender. We obtained the area under the receiver operating characteristic (AUROC), area under the precision-recall curve (AUPRC), true positive rate (TPR), positive predictive value (PPV), and negative predictive value (NPV). We report TPR, PPV and NPV by thresholding the algorithm output at a value that obtains a specificity of 90%. Based on previously published protocol for phenotyping studies, we defined the AD cohort at 90% specificity as a pragmatic decision to strike a balance between sensitivity and PPV while minimizing false positives.[15] We obtained the AUPRC in addition to the AUROC for all phenotyping algorithms as AUPRC is robust to sample size imbalance.[16] To assess the tradeoff between specificity and PPV, we also provided a comparison of the model performance at different specificity thresholds.

*S-Method-2.5. Creating AD cohorts for sensitivity analyses*

We conducted sensitivity analyses to ensure the robustness of our findings. We performed two sensitivity analyses: (1) by including a weighted comorbidity index score instead of individual comorbidities, and (2) by excluding patients with missing race and ethnicity information. In the first sensitivity analysis, we included a single score indicative of pre-existing comorbidity burden (Elixhauser comorbidity index with van Walraven weighting) instead of individual comorbidities.[17,18] In the second sensitivity analysis, we excluded patients with missing race and ethnicity information from the initial UPMC and MGB AD-EHR data-marts (*i.e., without* imputation of missing race and ethnicity) (**S-Figure 1**). Similar to the main AD cohorts, we created sensitivity analysis AD cohorts by including individuals predicted as having AD by KOMAP at 90% specificity. We excluded patients with <24 months of EHR data or admitted to nursing homes prior to the baseline.

**S-Method-3. Validation of nursing home admission status**

We randomly sampled 100 randomly selected patients at UPMC and MGB that met the study inclusion criteria (as shown in the **main Figure 1A**) for chart review of nursing home admission status. A clinical domain expert (SV) reviewed the EHR of these patients to determine nursing home admission status and used the annotated labels as the gold-standard ground truth for validating rule-based nursing home definitions from the EHR. We compared the performance of two definitions of nursing home admission: (1) ≥1 code for admission to or clinical encounters in any type of residential institution and (2) ≥1 code **or** CUI (narrative mention) for admission to **or** clinical encounters in any type of residential institution (**S-Table 3**).

#### S-Method-4. Statistical analysis

*S-Method-4.1. Baseline characteristics of the AD cohorts*

We used two-tailed t-tests to compare differences in demographics, baseline healthcare utilization, pre-existing comorbidity burden, prevalence of AD-related medication prescriptions (**S-Table 4**), and prevalence of outcomes of AD decline among demographic groups. We calculated the proportion of patients with each comorbidity in the ECI by demographic group and used two-proportion Z-tests to compare the difference in proportion of patients with each comorbidity among demographic groups.

*S-Method-4.2. Survival analysis and fixed-effects meta-analysis*

We estimated the time-to-outcome within each healthcare system using covariate-adjusted Cox proportional hazards (PH) models that accounted for the competing risks of nursing home admission and death, stratified by demographic groups. Using patient-level data from both sites, we then performed a *fixed*-effects meta-analysis of the competing risk Cox proportional hazard model of time to nursing home admission vs death across UPMC and MGB AD cohorts, as *random*-effects models would be highly unstable given the number of sites (*i.e.,* 2 healthcare systems). We used inverse variance weighting to combine the data from both sites. We estimated the time-to-outcome using healthcare system-*specific* (*i.e.,* at each healthcare system) covariate-adjusted competing risk Cox proportional hazards models stratified by demographic groups, with adjustment for the pooled covariates. We performed stratified analyses according to demographic groups by estimating the time-to-outcome for each patient while holding other covariates constant, except for the demographic variable of stratification (*i.e.,* gender, or race and ethnicity). This allowed for estimating the time-to-outcome for each demographic group, independent of the effect of additional covariates. We constructed covariate-adjusted survival curves stratified according to demographic groups. We obtained the adjusted hazard ratios for each covariate and visualized the fixed-effects meta-analysis results using forest plots.

#### S-Method-5. Data availability

We will publicly disseminate anonymous summary-level registry data and EHR data. The rationale for not sharing patient-level data is that patient-level clinical data (either de-identified information or limited protected health information containing dates of clinical events or even if anonymous due to concern for re-identification) are universally subject to the rules and regulation of each healthcare system, which may only be affiliated with but are not the same as the primary academic institutions of the study investigators. Sharing of de-identified EHR data with qualified external researchers by each of the study performance site may be permissible only after the approval of the respective Institutional Review Boards (IRBs), regulatory oversight agents of the healthcare systems (that own the clinical data) as well as the appropriate Data Usage Agreements (DUA) between institutions.

### SUPPLEMENTARY RESULTS

**S-Result-1. AD diagnosis phenotyping algorithm (KOMAP) performance**

The demographic profiles of the randomly selected patients for chart review at UPMC (64% women, 87% NHW, mean[SD]=81.55[9.00] years at AD diagnosis) and MGB (55% women, 90% NHW, mean[SD]=79.18[10.43] years at AD diagnosis) were similar to the larger UPMC (64% women, 92% NHW, mean[SD]=80.56[9.04] years at AD diagnosis) and MGB (59% women, 89% NHW, mean[SD]=78.63[9.58] years at AD diagnosis) AD-EHR cohorts (**S-Table 5**). Using chart-reviewed labels to evaluate performance, KOMAP achieved a higher AUROC at MGB (n=100, 45% AD, AUROC=0.923) than UPMC (n=200, 49% AD, AUROC=0.854) (**Figure 1B-E, S-Table 6, S-Table 7, S-Table 8, S-Table 9**). At 90% specificity, the sensitivity and PPV were reasonably balanced at both UPMC and MGB, similar to prior phenotyping studies (**S-Table 7**).[15] (Neither AUROC nor AUPRC changed when altering the specificity thresholds.) While KOMAP achieved higher AUROC in women at both healthcare systems (UPMC: women=0.888, men=0.782; MGB: women=0.965, men=0.855), there was inconsistent pattern between two systems for race and ethnicity groups (UPMC: NHW=0.856, other=0.854; MGB: NHW=0.925, other=1.000). AUPRC, which accounts for imbalance in the sample size, was higher among women than men at both UPMC (AUPRC: women=0.866, men=0.709) and MGB (AUPRC: women=0.926, men=0.868). There was an inconsistent pattern in AUPRC between two sites for race and ethnicity groups (UPMC: NHW=0.843, other=0.732; MGB: NHW=0.903, other=1.000). Using registry-derived labels to evaluate algorithm performance at UPMC (n=1916, 47% AD, AUROC=0.835, **S-Table 10**), KOMAP achieved higher AUROC and AUPRC in women than men and higher in racial and ethnic minorities than NHW individuals (AUROC: women=0.846, men=0.824; NHW=0.821, other=0.922; AUPRC: women=0.867, men=0.784; NHW=0.824, other=0.906) respectively.

KOMAP performance evaluated with both gold-standard chart-reviewed and registry-derived labels outperformed all the benchmark methods constructed from surrogate features of AD diagnosis in terms of AUROC and AUPRC, demonstrating the advantage of incorporating information from knowledge graph-selected features using KOMAP (**S-Table 8, S-Table 9, S-Table 10**). Training KOMAP separately for each demographic group (KOMAP separate) resulted in similar performance. KOMAP performance in the sensitivity analysis AD cohorts (without imputation of missing race and ethnicity) evaluated using chart-reviewed labels was consistent with the main AD cohorts (**S-Table 11**).

**S-Result-2. Validation of nursing home admission status**

The rule-based definition of ≥1 code *or* CUI for nursing home admission outperformed ≥1 code for nursing home admission at both UPMC (≥1 code: sensitivity=0.493, specificity=0.968, PPV=0.971, NPV=0.462; ≥1 code *or* CUI: sensitivity=0.913, specificity=0.903, PPV=0.955, NPV=0.824, **S-Table 12**) and MGB (≥1 code: sensitivity=0.709, specificity=1, PPV=1, NPV=0.738; ≥1 code or CUI: sensitivity=0.945, specificity=0.911, PPV=0.929, NPV=0.932). The definition of '≥1 code *or* CUI' greatly increased the sensitivity and NPV over the definition of '≥1 code', while slightly decreasing the specificity and PPV.

**S-Result-3. AD cohort characteristics for sensitivity analyses**

In the first sensitivity analysis using the van Walraven weighted Elixhauser score (instead of individual comorbidities) as a covariate, we included all patients from the main combined AD cohort (*with* imputation of missing race and ethnicity, **main Table 1**).

In the second sensitivity analysis, the combined AD cohort (*without* imputation of missing race and ethnicity) comprised 24,075 patients (61% women, 90% NHW, mean[SD]=79.55[9.45] years at AD diagnosis, **S-Table 13**), including 12,938 UPMC patients (64% women, 92% NHW, 80.58[9.01] years at AD diagnosis) and 11,137 MGB patients (59% women, 88% NHW, 78.35[9.81] years at AD diagnosis). Similar to the main AD cohorts, when compared to MGB patients, UPMC patients had older age at AD diagnosis, greater proportion of women and NHW individuals, lower baseline healthcare utilization, greater pre-existing comorbidity burden (**S-Figure 2**), greater proportion of nursing home admission and death during study follow-up, and higher proportion of AD-related medication prescriptions (particularly before AD diagnosis).

**S-Result-4. Sensitivity analyses of the differential risk of nursing home admission and death**

For sensitivity analyses, we again used fixed-effects meta-analysis of covariate-adjusted Cox proportional hazard models to compare the risk of two readily ascertainable clinical outcomes among demographic groups.

In the first sensitivity analysis when using the van Walraven weighted Elixhauser score as covariate, the results were consistent results with the main findings. Women had a higher risk of nursing home admission than men (HR [95% CI]=1.084 [1.047-1.122], *p*<.0001, **main Figure 3, S-Table 14, S-Figure 3**, **S-Figure 4**), but there was no significant difference between racial and ethnic groups (HR [95% CI]=0.962 [0.912-1.016], *p*=.163). Women had a lower risk of death than men and NHW individuals had a higher risk of death than racial and ethnic minorities (women HR [95% CI]=0.860 [0.816-0.907], *p*<.0001; NHW HR [95% CI]=1.360 [1.233-1.501], *p*<.0001). Consistent with the main findings, higher Elixhauser scores (indicative of greater cumulative pre-existing comorbidity burden) were associated with increased risk of both nursing home admission and death (nursing home admission HR [95% CI]=1.008 [1.006-1.010], *p*<.0001; death HR [95% CI]=1.026 [1.023-1.030], *p*<.0001, **main Figure 4**, **S-Figure 4**).

In the second sensitivity analysis, the combined AD cohort (*without* imputation of missing race and ethnicity) showed consistent results as the main combined AD cohort (*with* imputation of missing race and ethnicity). Women had a higher risk of nursing home admission than men (HR [95% CI]=1.065 [1.025-1.107], *p*=.001, **main Figure 3, S-Table 14**, **S-Figure 5**, **S-Figure 6**), but there was no significant difference between racial and ethnic groups (HR [95% CI]=1.009 [0.952-1.069], *p*=.767). Women had a lower risk of death than men and NHW individuals had a higher risk of death than racial and ethnic minorities (women HR [95% CI]=0.823 [0.769-0.882], *p*<.0001; NHW HR [95% CI]=1.458 [1.283-1.657], *p*<.0001, **main Figure 3**, **S-Table 15**, **S-Figure 5**, **S-Figure 6**). Consistent with the main combined AD cohort (*with* imputation of missing race and ethnicity), the key drivers of nursing home admission and death risk in the combined sensitivity analysis AD cohort (*without* imputation of missing race and ethnicity) included older age at AD diagnosis and greater pre-existing comorbidity burden (**S-Figure 6**).

### SUPPLEMENTARY DISCUSSION

**S-Discussion-1. Differences in AD diagnosis phenotyping algorithm (KOMAP) performance across healthcare systems and demographic groups**

KOMAP demonstrated significantly better performance than rule-based methods, outperforming all the benchmark methods based on surrogate features of AD diagnosis (i.e., main AD PheCode, main dementia PheCode, main AD CUI). Specifically, the performance of the rule-based approaches in this study using surrogate features was comparable to previously published studies of AD cohort identification.[3,4,10] KOMAP, while performing well overall, showed modest differences in healthcare system- and demographic group-specific performances, all of which still achieved reasonable AUROCs and AURPCs. To avoid co-training, we trained KOMAP independently at UPMC and MGB. KOMAP performed better at MGB compared to UPMC, potentially due to differences in clinical practices, documentation standards, population characteristics, and EHR data quality between the two healthcare systems. At both healthcare systems, KOMAP performed better among women than men, possibly attributable to women having more available EHR data for algorithm than men for several related reasons: (1) women had higher incidence and prevalence of AD;[19–23] (2) the real-world AD cohorts had older age at AD diagnosis than traditional epidemiological studies;[24,25] (3) women survived longer than men, including after AD diagnosis.[26,27] Men with AD might also have greater overlap with other related dementias.[28] There was no consistent pattern of performance differences by race and ethnicity. KOMAP performed equally well among racial and ethnic groups at UPMC, while KOMAP in racial and ethnic minorities outperformed NHW individuals at MGB. The different patterns of healthcare utilization between UPMC and MGB populations might be contributory. KOMAP deployment for AD diagnosis in future healthcare systems will require healthcare system-specific validation. Despite modest algorithm performance differences, the final combined AD cohort will be valuable for generating real-world evidence, particularly across demographic groups.

**S-Discussion-2. Innovation of the AD diagnosis phenotyping approach (KOMAP)**

KOMAP had the following innovations that enhanced its application when compared to prior methods.[3–8,10–12,22,29,30] First, most prior AD diagnosis phenotyping algorithms relied on literature review or expert curation of features, which limited scalability. In contrast, KOMAP leveraged pre-trained and publicly available knowledge graphs of interconnected EHR concepts to rapidly select informative features, a strategy superior to reliance on clinician experts while effectively reducing feature dimensionality. Second, to validate the AD diagnosis phenotyping algorithms, prior studies used labels such as clinical assessment, chart review, and surrogate indicators within the EHR. KOMAP did not require gold-standard labels for training, though we used gold-standard labels to provide a highly reliable estimate of algorithm performance when available.

### SUPPLEMENTARY TABLES

**S-Table 1.** Initial AD data mart specifications.

| Code | Description |
| --- | --- |
| ICD-9 290.0 | Senile dementia, uncomplicated |
| ICD-9 290.1 | Presenile dementia |
| ICD-9 290.10 | Presenile dementia, uncomplicated |
| ICD-9 290.11 | Presenile dementia with delirium |
| ICD-9 290.12 | Presenile dementia with delusional features |
| ICD-9 290.13 | Presenile dementia with depressive features |
| ICD-9 290.20 | Senile dementia with delusional features |
| ICD-9 290.21 | Senile dementia with depressive features |
| ICD-9 290.3 | Senile dementia with delirium |
| ICD-9 294.20 | Dementia, unspecified, without behavioral disturbance |
| ICD-9 294.21 | Dementia, unspecified, with behavioral disturbance |
| ICD-9 331.0 | Alzheimer’s disease |
| ICD-10 F03.90 | Unspecified dementia without behavioral disturbance |
| ICD-10 F03.91 | Unspecified dementia with behavioral disturbance |
| ICD-10 G30 | Alzheimer's disease |
| ICD-10 G30.0 | Alzheimer's disease with early onset |
| ICD-10 G30.1 | Alzheimer's disease with late onset |
| ICD-10 G30.8 | Other Alzheimer's disease |
| ICD-10 G30.9 | Alzheimer's disease, unspecified |

Note: Inclusion criteria for the initial AD data mart were having ≥1 ICD code for AD or related dementia.

**S-Table 2.** Coefficients of features in AD diagnosis phenotyping algorithm.

| Code/CUI | Term | Coefficient [a] |
|---|---|---|
| PheCode.290.11 | Alzheimer's disease | 0.5886 |
| C0002395 | Alzheimer's disease | 0.6073 |
| PheCode.290 | Delirium dementia and amnestic and other cognitive disorders | 0.0362 |
| PheCode.290.1 | Dementias | 0.0530 |
| RXNORM.135447 | Donepezil | 0.0072 |
| RXNORM.6719 | Memantine | 0.0111 |
| RXNORM.183379 | Rivastigmine | 0.0037 |
| PheCode.292 | Neurological disorders | -0.0041 |
| RXNORM.4637 | Galantamine | 0.0000 |
| PheCode.433 | Cerebrovascular disease | -0.0035 |
| PheCode.292.4 | Altered mental status | -0.0060 |
| PheCode.332 | Parkinson's disease | 0.0000 |
| PheCode.350.2 | Abnormality of gait | -0.0018 |
| RXNORM.103990 | Carbidopa/levodopa | -0.0021 |
| RXNORM.1791685 | Pimavanserin | 0.0000 |
| RXNORM.25025 | Finasteride | 0.0000 |
| RXNORM.51272 | Quetiapine | 0.0000 |
| RXNORM.77492 | Tamsulosin | -0.0011 |
| RXNORM.11248 | Cyanocobalamin | 0.0000 |
| RXNORM.15996 | Mirtazapine | 0.0000 |
| PheCode.433.8 | Late effects of cerebrovascular disease | 0.0000 |
| PheCode.333.3 | Tics and choreas | 0.0000 |
| RXNORM.61381 | Olanzapine | 0.0000 |
| RXNORM.36437 | Sertraline | 0.0000 |
| PheCode.295 | Schizophrenia and other psychotic disorders | -0.0001 |
| RXNORM.35636 | Risperidone | 0.0000 |
| RXNORM.114477 | Levetiracetam | 0.0000 |
| RXNORM.43611 | Latanoprost | 0.0000 |
| PheCode.433.3 | Cerebral ischemia | 0.0000 |
| RXNORM.17767 | Amlodipine | -0.0021 |
| PheCode.433.21 | Cerebral artery occlusion, with cerebral infarction | -0.0007 |
| RXNORM.221147 | Polyethylene glycol 3350 | -0.0024 |
| PheCode.798 | Malaise and fatigue | -0.0049 |
| PheCode.591 | Urinary tract infection | -0.0020 |
| PheCode.430.2 | Intracerebral hemorrhage | 0.0000 |
| PheCode.818 | Intracranial hemorrhage (injury) | 0.0000 |
| PheCode.345 | Epilepsy, recurrent seizures, convulsions | 0.0000 |
| PheCode.430 | Intracranial hemorrhage | 0.0000 |
| RXNORM.321988 | Escitalopram | 0.0000 |
| RXNORM.10737 | Trazodone | 0.0000 |
| C0011265 | Presenile dementia | 0.0231 |
| C0497327 | Dementia | 0.0234 |

| | | |
|---|---|---|
| C0527316 | Donepezil | 0.0110 |
| C0011269 | Dementia, vascular | -0.0065 |
| C0025242 | Memantine | 0.0118 |
| C0085220 | Cerebral amyloid angiopathy | 0.0000 |
| C3714756 | Intellectual disability | 0.0000 |
| C0034770 | Mental recall | -0.0003 |
| C0242422 | Parkinsonian disorders | 0.0000 |
| C4084203 | Improved - answer to question | -0.0028 |
| C0009676 | Confusion | 0.0027 |
| C0003537 | Aphasia | 0.0000 |
| C0751587 | CADASIL syndrome | 0.0000 |
| C4521042 | Complete trisomy 21 syndrome | 0.0000 |
| C0030567 | Parkinson disease | -0.0007 |
| C3531686 | Ginkgo biloba whole | 0.0000 |
| C0004093 | Asthenia | -0.0011 |
| C0524188 | Methylphenidate measurement | 0.0000 |
| C5201148 | Moderate | -0.0001 |
| C3714552 | Weakness | -0.0015 |
| C0439044 | Living alone | -0.0022 |
| C0085639 | Falls | -0.0005 |
| C0018786 | Hearing tests | -0.0003 |
| C0595998 | Household composition | 0.0016 |
| C0518460 | Bathing self care | 0.0000 |
| C1548428 | Referral type - Psychiatric | -0.0017 |
| C0260942 | Encounter due to screening for depression | 0.0000 |
| C3526598 | Psychiatric service | -0.0015 |
| C0038454 | Cerebrovascular accident | -0.0018 |
| C0430533 | Dental diagnostic procedure | 0.0000 |
| C4760315 | Obsessive compulsive disorder drugs | 0.0000 |
| C4759845 | CTCAE v4 Grade 2 | -0.0001 |
| C0374711 | Surgical repair | 0.0000 |
| C3537736 | Once a day | -0.0010 |
| C1384666 | Hearing impairment | 0.0000 |
| C3811652 | A proliferation-inducing ligand measurement | 0.0000 |
| C0168634 | Baseline dental cement | 0.0004 |
| C4534363 | At home | -0.0005 |
| C1266533 | Fluvoxamine measurement | 0.0000 |
| C2936883 | Vitamin B12 [EPC] | 0.0016 |
| C0007787 | Transient ischemic attack | 0.0017 |
| C0042845 | Vitamin B 12 | 0.0015 |
| C0034991 | Rehabilitation therapy | -0.0029 |
| C0011127 | Pressure ulcer | 0.0000 |
| C0011053 | Deafness | 0.0000 |
| C0008845 | Citalopram | 0.0002 |
| C0757844 | TNFSF13 protein, human | 0.0000 |

| | | |
|---|---|---|
| C0018772 | Hearing loss, partial | 0.0000 |
| C0042571 | Vertigo | 0.0000 |
| C3809991 | Immunodeficiency, common variable, 10 | 0.0000 |
| C1262477 | Weight decreased | 0.0000 |
| C5441521 | Complaint (finding) | 0.0000 |
| C0007820 | Cerebrovascular disorders | 0.0000 |
| C0425245 | Mobility as a finding | 0.0000 |
| C0011168 | Deglutition disorders | 0.0000 |
| C1522133 | Hypercholesterolemia result | 0.0000 |
| C4553189 | Citalopram measurement | 0.0002 |
| C0428977 | Bradycardia | 0.0000 |
| C4484264 | Clark level | 0.0000 |
| C3714760 | Drug-induced tardive dyskinesia | 0.0000 |
| C5399953 | Methotrexate drug assay | 0.0000 |
| C4763868 | Cocaine use disorder | 0.0000 |
| C0202385 | Fluoxetine measurement | 0.0000 |
| C1847835 | Vitiligo-associated multiple autoimmune disease susceptibility 1 | 0.0000 |
| C4042877 | Clinical decision-making | 0.0013 |
| C0123091 | Quetiapine | 0.0000 |
| C0373527 | Acetaminophen assay | 0.0000 |
| C2917659 | Acetaminophen [EPC] | 0.0000 |
| C4522247 | Allogeneic blood product donation | 0.0000 |
| C0206275 | Widowhood | 0.0000 |
| C4317146 | Acid reflux | 0.0000 |
| C3853703 | Pacemaker ECG assessment | 0.0000 |
| C1880851 | Fracture of medical device material | 0.0000 |
| C2266644 | Subjective (symptom) | 0.0000 |
| C0278061 | Abnormal mental state | 0.0007 |
| C3683798 | Graft procedures on the head | 0.0000 |
| C4721453 | Peripheral nervous system diseases | 0.0000 |
| C0042029 | Urinary tract infection | 0.0060 |
| C0022983 | Laminectomy | 0.0000 |
| C0202361 | Clonazepam measurement | 0.0000 |
| C4551529 | Renal dialysis | 0.0000 |
| C1332206 | Adult lymphoma | 0.0000 |
| C0041296 | Tuberculosis | 0.0000 |
| C5425799 | All other | 0.0000 |
| C0019360 | Herpes zoster disease | 0.0000 |
| C2015933 | Outcomes otolaryngology hearing | -0.0001 |
| C1856053 | Hydranencephaly with renal aplasia-dysplasia | 0.0000 |
| C3662068 | Static encephalopathy | 0.0000 |
| C2227886 | PSEN1 gene (procedure) | 0.0000 |
| C2174421 | Examination of balance | -0.0039 |
| C1551394 | Normal device alert level | 0.0000 |
| C0393483 | Brainstem encephalitis | 0.0000 |

| | | |
|---|---|---|
| C0149843 | Punch drunk syndrome | 0.0000 |
| C0751438 | Posterior pituitary disease | 0.0000 |
| C2227885 | APP gene (procedure) | 0.0000 |
| C0238111 | Lennox-Gastaut syndrome | 0.0000 |
| C3835651 | Resident - answer to question | -0.0030 |
| C4048351 | Galactocerebrosidase, human | 0.0000 |
| C1873497 | Normal assessment finding | -0.0006 |
| C1550457 | Normal observation interpretation | -0.0009 |
| C4551993 | Amyotrophic lateral sclerosis, familial | 0.0000 |
| C1561581 | Allergy severity - severe | 0.0000 |
| C0728873 | Monitor brand of insecticide | 0.0000 |
| C4722630 | Solution dosage form category | -0.0001 |
| C0016957 | Galactosylceramidase | 0.0000 |
| C0424653 | Weight symptom (finding) | 0.0046 |

a. Coefficient indicates the importance of the feature for the target disease (*i.e.,* AD). Features with positive coefficients are more closely related to the target disease, whereas features with negative coefficients are less related to the target disease.

**S-Table 3.** List of codes and concept unique identifiers (CUIs) for nursing home admission.

| Code | Description |
|---|---|
| ICD9CM:E849.7 | Accidents occurring in residential institution |
| ICD9CM:V60.6 | Person living in residential institution |
| ICD9CM:V63.2 | Person awaiting admission to adequate facility elsewhere |
| ICD9CM:V66.7 | Encounter for palliative care |
| ICD10:Y92.120 | Kitchen in nursing home as the place of occurrence of the external cause |
| ICD10:Y92.121 | Bathroom in nursing home as the place of occurrence of the external cause |
| ICD10:Y92.122 | Bedroom in nursing home as the place of occurrence of the external cause |
| ICD10:Y92.123 | Driveway of nursing home as the place of occurrence of the external cause |
| ICD10:Y92.128 | Other place in nursing home as the place of occurrence of the external cause |
| ICD10:Y92.129 | Unspecified place in nursing home as the place of occurrence of the external cause |
| ICD10CM:Z02.2 | Encounter for examination for admission to residential institution |
| ICD10CM:Z51.5 | Encounter for palliative care |
| ICD10CM:Z59.3 | Problems related to living in residential institution |
| ICD10CM:Z74.1 | Problems related to care provider dependency |
| ICD10CM:Z75.1 | Person awaiting admission to adequate facility elsewhere |
| CPT4:99301 | Evaluation and management of a new or established patient involving an annual nursing facility assessment: 30 minutes at the bedside |
| CPT4:99302 | Evaluation and management of a new or established patient involving an annual nursing facility assessment of a complication or a new problem: 40 minutes at the bedside |
| CPT4:99303 | Evaluation and management of a new or established patient involving an annual nursing facility assessment at the time of initial admission to the facility: 50 minutes at the bedside |
| CPT4:99304 | Initial nursing facility care 25 min |
| CPT4:99305 | Initial nursing facility care 35 min |
| CPT4:99306 | Initial nursing facility care 45 min |
| CPT4:99307 | Subsequent nursing facility care, per day, for the evaluation and management of a patient: 10 minutes at the bedside |
| CPT4:99308 | Subsequent nursing facility care, per day, for the evaluation and management of a patient: 15 minutes at the bedside |
| CPT4:99309 | Subsequent nursing facility care, per day, for the evaluation and management of a patient: 25 minutes at the bedside |
| CPT4:99310 | Subsequent nursing facility care, per day, for the evaluation and management of a patient: 35 minutes at the bedside |

| | |
|---|---|
| CPT4:99311 | Subsequent nursing facility care, per day, for the evaluation and management of a new or established patient: 15 minutes at the bedside |
| CPT4:99312 | Subsequent nursing facility care, per day, for the evaluation and management of a new or established patient who is responding inadequately to therapy or has developed a minor complication: 25 minutes at the bedside |
| CPT4:99313 | Subsequent nursing facility care, per day, for the evaluation and management of a new or established patient who has developed a significant complication or a new problem: 35 minutes at the bedside |
| CPT4:99315 | Nursing facility discharge day management; 30 minutes or less |
| CPT4:99316 | Nursing facility discharge day management; more than 30 minutes |
| CPT4:99318 | Evaluation and management of a patient involving an annual nursing facility assessment |
| CPT4:99377 | Report this service when the provider supervises and coordinates the care provided to a hospice patient. |
| CPT4:99378 | Report this service when the provider supervises and coordinates the care provided to a hospice patient. |
| CPT4:99379 | Physician supervision of a nursing facility patient (patient not present) requiring complex and multidisciplinary care; 15–29 minutes |
| CPT4:99380 | Physician supervision of a nursing facility patient (patient not present) requiring complex and multidisciplinary care modalities; 30 minutes or more |
| HCPCS:G0066 | Physician supervision of a nursing facility patient (patient not present); 30 minutes or more per month |
| HCPCS:G0299 | Direct skilled nursing services of a registered nurse (RN) in the home health or hospice setting, each 15 minutes |
| HCPCS:G0300 | Direct skilled nursing services of a licensed practical nurse (LPN) in the home health or hospice setting, each 15 minutes |
| HCPCS:G9685 | Physician service or other qualified health care professional for the evaluation and management of a beneficiary's acute change in condition in a nursing facility |
| HCPCS:Q5001 | Hospice or home health care provided in patient's home/residence |
| HCPCS:Q5002 | Hospice or home health care provided in assisted living facility |
| HCPCS:Q5003 | Hospice care provided in nursing long term care facility (LTC) or non-skilled nursing facility (NF) |
| HCPCS:Q5004 | Hospice care provided in skilled nursing facility (SNF) |
| HCPCS:Q5005 | Hospice care provided in inpatient hospital |
| HCPCS:Q5006 | Hospice care provided in inpatient hospice facility |
| HCPCS:Q5007 | Hospice care provided in long term care facility |
| HCPCS:Q5008 | Hospice care provided in inpatient psychiatric facility |

| | |
|---|---|
| HCPCS:Q5009 | Hospice or home health care provided in place not otherwise specified (NOS) |
| HCPCS:Q5010 | Hospice home care provided in a hospice facility |
| HCPCS:T2042 | Hospice routine home care per diem |
| HCPCS:T2043 | Hospice continuous home care per hour |
| HCPCS:T2044 | Hospice inpatient respite care per diem |
| HCPCS:T2045 | Hospice general inpatient care per diem |
| HCPCS:T2046 | Hospice long term care, room and board only per diem |
| C0028688 | Nursing home |
| C0037265 | Nursing home |
| C0184696 | Nursing home |
| C0260062 | Nursing home |
| C0338041 | Nursing home |
| C0402670 | Nursing home |
| C0419370 | Nursing home |
| C0421614 | Nursing home |
| C0422260 | Nursing home |
| C0425205 | Nursing home |
| C0500310 | Nursing home |
| C0500311 | Nursing home |
| C0545083 | Nursing home |
| C0557820 | Nursing home |
| C0580114 | Nursing home |
| C0584536 | Nursing home |
| C0584537 | Nursing home |
| C0587730 | Nursing home |
| C0682287 | Nursing home |
| C0741955 | Nursing home |
| C0741956 | Nursing home |
| C0741957 | Nursing home |
| C0741958 | Nursing home |
| C0848616 | Nursing home |
| C0850472 | Nursing home |
| C0850480 | Nursing home |
| C0850485 | Nursing home |
| C0850570 | Nursing home |
| C1266870 | Nursing home |
| C1517936 | Nursing home |
| C1553047 | Nursing home |
| C1657223 | Nursing home |
| C1658399 | Nursing home |

| | |
|---|---|
| C1714469 | Nursing home |
| C1714474 | Nursing home |
| C1717017 | Nursing home |
| C1717037 | Nursing home |
| C1717654 | Nursing home |
| C1717730 | Nursing home |
| C1717899 | Nursing home |
| C1719075 | Nursing home |
| C1830397 | Nursing home |
| C2046796 | Nursing home |
| C2081544 | Nursing home |
| C2108009 | Nursing home |
| C2193542 | Nursing home |
| C2230060 | Nursing home |
| C2585549 | Nursing home |
| C2707434 | Nursing home |
| C2707708 | Nursing home |
| C2907705 | Nursing home |
| C2907706 | Nursing home |
| C2907707 | Nursing home |
| C2907708 | Nursing home |
| C2907709 | Nursing home |
| C2907710 | Nursing home |
| C2907711 | Nursing home |
| C2907712 | Nursing home |
| C2907713 | Nursing home |
| C2907714 | Nursing home |
| C2960681 | Nursing home |
| C3530270 | Nursing home |
| C3640848 | Nursing home |
| C3648882 | Nursing home |
| C3650604 | Nursing home |
| C3714457 | Nursing home |
| C3840746 | Nursing home |
| C3841564 | Nursing home |
| C3844657 | Nursing home |
| C3844664 | Nursing home |
| C3844665 | Nursing home |
| C3844666 | Nursing home |
| C3844667 | Nursing home |
| C3844668 | Nursing home |

| | |
|---|---|
| C3844669 | Nursing home |
| C3844670 | Nursing home |
| C3844671 | Nursing home |
| C3844765 | Nursing home |
| C3845163 | Nursing home |
| C3845568 | Nursing home |
| C3871304 | Nursing home |
| C4047960 | Nursing home |
| C4081949 | Nursing home |
| C4264552 | Nursing home |
| C4534419 | Nursing home |
| C4534566 | Nursing home |
| C4535882 | Nursing home |
| C4698451 | Nursing home |
| C4746466 | Nursing home |
| C5419144 | Nursing home |
| C5437159 | Nursing home |
| C5452855 | Nursing home |
| C5452867 | Nursing home |

**S-Table 4.** List of AD-related medications.

| Brand Name | Generic Name | Treatment Class |
|---|---|---|
| Adlarity | Donepezil | Acetylcholinesterase Inhibitor |
| Aricept | Donepezil | Acetylcholinesterase Inhibitor |
| Exelon | Rivastigmine | Acetylcholinesterase Inhibitor |
| Razadyne ER | Galantamine | Acetylcholinesterase Inhibitor |
| Razadyne [DSC] | Galantamine | Acetylcholinesterase Inhibitor |
| Namenda | Memantine | N-Methyl-D-Aspartate (NMDA) Receptor Antagonist |
| Namenda Titration Pak | Memantine | N-Methyl-D-Aspartate (NMDA) Receptor Antagonist |
| Namenda XR | Memantine | N-Methyl-D-Aspartate (NMDA) Receptor Antagonist |
| Namenda XR Titration Pack [DSC] | Memantine | N-Methyl-D-Aspartate (NMDA) Receptor Antagonist |
| Aduhelm | Aducanumab | Anti-Amyloid Monoclonal Antibody |
| Leqembi | Lecanemamb-irmb | Anti-Amyloid Monoclonal Antibody |
| Namzaric | Donepezil and memantine | Acetylcholinesterase Inhibitor; N-Methyl-D-Aspartate (NMDA) Receptor Antagonist |

**S-Table 5.** Demographic and clinical profile of the patients randomly selected for chart review for AD diagnosis at UPMC and MGB.

| | **University of Pittsburgh ADRC registry** | **UPMC chart review** | **MGB chart review** | **P-value [a]** |
|---|---|---|---|---|
| **Number of Patients, N (%)** | 1916 (100%) | 200 (100%) | 100 (100%) | |
| **Number of AD Patients, N (%)** | 904 (47%) | 98 (49%) | 45 (45%) | |
| **Age at AD diagnosis [b], mean (SD)** | 73.88 (8.93) | 81.55 (9.00) | 79.18 (10.43) | 0.2 |
| **Gender, N (%)** | | | | 0.2 |
| Women | 1120 (58%) | 127 (64%) | 55 (55%) | |
| Men | 796 (42%) | 73 (37%) | 45 (45%) | |
| **Race and ethnicity, N (%)** | | | | |
| Non-Hispanic White | | 173 (87%) | 90 (90%) | 0.2 |
| American Indian | 1676 (87%) | <10 (<0.1%) | 0 (0%) | |
| Asian | <10 (<0.1%) | <10 (<0.1%) | <10 (<0.1%) | |
| Black | 14 (0.7%) | 24 (12%) | <10 (<0.1%) | |
| Hawaiian or Pacific Islander | 216 (11%) | 0 (0%) | <10 (<0.1%) | |
| Hispanic or Latino | 0 (0%) | <10 (<0.1%) | <10 (<0.1%) | |
| Other | <10 (<0.1%) | 0 (0%) | <10 (<0.1%) | |

a. P-value obtained from a two-tailed t-test comparing UPMC and MGB chart review label data.
b. Age reported only for AD patients.

**S-Table 6.** Performance of the AD diagnosis phenotyping algorithm (held at 90% specificity) evaluated using chart-reviewed labels at UPMC and MGB across demographic groups.

| | UPMC (n = 200) | | | | | MGB (n = 100) | | | | |
|---|---|---|---|---|---|---|---|---|---|---|
| | **All** | **NHW [b]** | **Other [c]** | **Men** | **Women** | **All** | **NHW [b]** | **Other [c]** | **Men** | **Women** |
| **AUROC** | 0.854 | 0.858 | 0.854 | 0.782 | 0.888 | 0.923 | 0.925 | 1.000 | 0.855 | 0.965 |
| **AUPRC** | 0.825 | 0.843 | 0.732 | 0.709 | 0.866 | 0.904 | 0.903 | 1.000 | 0.868 | 0.926 |
| **Sensitivity [a]** | 0.595 | 0.602 | 0.630 | 0.447 | 0.726 | 0.750 | 0.857 | 1.000 | 0.667 | 0.808 |
| **PPV [a]** | 0.839 | 0.849 | 0.786 | 0.768 | 0.865 | 0.868 | 0.889 | 1.000 | 0.857 | 0.913 |
| **NPV [a]** | 0.716 | 0.807 | 0.709 | 0.687 | 0.789 | 0.804 | 0.875 | 1.000 | 0.778 | 0.833 |

a. We calculated the sensitivity, positive predictive value (PPV), and negative predictive value (NPV) at a pre-defined specificity of 90%. (See Methods for rationale of holding specificity at 90%.)
b. NHW, non-Hispanic White individuals
c. Other, racial and ethnic minorities

Other Abbreviations: *AUPRC*, area under the precision-recall curve; *AUROC,* area under the receiver operating characteristic curve; *NPV,* negative predictive value; *PPV*, positive predictive value.

**S-Table 7.** Performance of the AD diagnosis phenotyping algorithm evaluated using chart-reviewed labels at UPMC and MGB at different specificity thresholds.

| Site | Specificity Threshold | AUROC | AUPRC | Sensitivity | PPV | NPV |
|---|---|---|---|---|---|---|
| **UPMC** | 0.95 | 0.865 | 0.837 | 0.439 | 0.896 | 0.638 |
| | 0.90 | 0.865 | 0.837 | 0.612 | 0.857 | 0.708 |
| | 0.85 | 0.865 | 0.837 | 0.714 | 0.824 | 0.757 |
| | 0.80 | 0.865 | 0.837 | 0.765 | 0.789 | 0.781 |
| | 0.75 | 0.865 | 0.837 | 0.786 | 0.755 | 0.786 |
| **MGB** | 0.95 | 0.923 | 0.904 | 0.682 | 0.938 | 0.774 |
| | 0.90 | 0.923 | 0.904 | 0.750 | 0.868 | 0.804 |
| | 0.85 | 0.923 | 0.904 | 0.773 | 0.829 | 0.811 |
| | 0.80 | 0.923 | 0.904 | 0.863 | 0.792 | 0.870 |
| | 0.75 | 0.923 | 0.904 | 0.955 | 0.778 | 0.950 |

Abbreviations: *AUPRC*, area under the precision-recall curve; *AUROC,* area under the receiver operating characteristic curve; *NPV,* negative predictive value; *PPV*, positive predictive value.

**S-Table 8.** Comparative performance AD diagnosis phenotyping algorithm (held at 90% specificity) against benchmarks at UPMC evaluated using chart-reviewed labels.

| | **Method** | **All** | **Validation by Race and Ethnicity** [a] | | **Validation by Gender** | |
|---|---|---|---|---|---|---|
| | | | **NHW** [b] | **Other** [c] | **Men** | **Women** |
| AUROC | PheCode290.11 [d] | 0.816 | 0.817 | 0.827 | 0.724 | 0.871 |
| | PheCode290.1 [d] | 0.751 | 0.740 | 0.838 | 0.632 | 0.796 |
| | PheCode290 [d] | 0.743 | 0.736 | 0.820 | 0.627 | 0.793 |
| | C0002395 [d] | 0.831 | 0.832 | 0.838 | 0.767 | 0.855 |
| | **KOMAP** | 0.854 | 0.858 | 0.854 | 0.782 | 0.888 |
| | KOMAP separate [e] | 0.854 | 0.859 | 0.838 | 0.784 | 0.888 |
| AUPRC | PheCode290.11 | 0.769 | 0.793 | 0.679 | 0.658 | 0.829 |
| | PheCode290.1 | 0.658 | 0.651 | 0.680 | 0.524 | 0.730 |
| | PheCode290 | 0.651 | 0.648 | 0.665 | 0.508 | 0.729 |
| | C0002395 | 0.805 | 0.810 | 0.811 | 0.745 | 0.820 |
| | **KOMAP** | 0.825 | 0.843 | 0.732 | 0.709 | 0.866 |
| | KOMAP separate | 0.819 | 0.842 | 0.711 | 0.709 | 0.866 |
| TPR (at FPR≤ 0.1) | PheCode290.11 | 0.537 | 0.542 | 0.508 | 0.356 | 0.636 |
| | PheCode290.1 | 0.325 | 0.291 | 0.483 | 0.160 | 0.368 |
| | PheCode290 | 0.319 | 0.289 | 0.476 | 0.159 | 0.375 |
| | C0002395 | 0.592 | 0.603 | 0.739 | 0.441 | 0.627 |
| | **KOMAP** | 0.595 | 0.602 | 0.630 | 0.447 | 0.726 |
| | KOMAP separate | 0.601 | 0.603 | 0.634 | 0.446 | 0.727 |
| PPV (at FPR≤ 0.1) | PheCode290.11 | 0.815 | 0.824 | 0.754 | 0.734 | 0.842 |
| | PheCode290.1 | 0.726 | 0.715 | 0.714 | 0.536 | 0.774 |
| | PheCode290 | 0.726 | 0.718 | 0.706 | 0.528 | 0.779 |
| | C0002395 | 0.826 | 0.837 | 0.787 | 0.758 | 0.844 |
| | **KOMAP** | 0.839 | 0.849 | 0.786 | 0.768 | 0.865 |
| | KOMAP separate | 0.823 | 0.833 | 0.773 | 0.766 | 0.866 |
| NPV (at FPR≤ 0.1) | PheCode290.11 | 0.703 | 0.695 | 0.752 | 0.643 | 0.747 |
| | PheCode290.1 | 0.621 | 0.595 | 0.771 | 0.597 | 0.604 |
| | PheCode290 | 0.615 | 0.589 | 0.773 | 0.603 | 0.606 |
| | C0002395 | 0.733 | 0.727 | 0.873 | 0.694 | 0.736 |
| | **KOMAP** | 0.716 | 0.709 | 0.807 | 0.687 | 0.789 |
| | KOMAP separate | 0.697 | 0.688 | 0.779 | 0.689 | 0.788 |

a. We imputed missing race and ethnicity using multiple imputation based on age and gender.
b. NHW, non-Hispanic White individuals
c. Other, racial and ethnic minorities
d. Refer to S-Table 2 for explanation of PheCodes or Concept Unique Identifiers as benchmark.
e. “KOMAP separate” indicates that KOMAP was trained separately by race and ethnicity and by gender.

Other Abbreviations: *AUPRC*, area under the precision-recall curve; *AUROC,* area under the receiver operating characteristic curve; *FPR,* true positive rate; *NPV,* negative predictive value; *PPV*, positive predictive value; *TPR,* true positive rate.

**S-Table 9.** Comparative performance of AD diagnosis phenotyping algorithm (held at 90% specificity) against benchmarks at MGB evaluated using chart-reviewed labels.

| | **Method** | **All** | **Validation by Race and Ethnicity [a]** | | **Validation by Gender** | |
|---|---|---|---|---|---|---|
| | | | **NHW [b]** | **Other [c]** | **Men** | **Women** |
| AUROC | PheCode290.11 [d] | 0.843 | 0.867 | 0.667 | 0.831 | 0.852 |
| | PheCode290.1 [d] | 0.735 | 0.732 | 0.933 | 0.634 | 0.803 |
| | PheCode290 [d] | 0.684 | 0.705 | 0.700 | 0.593 | 0.741 |
| | C0002395 [d] | 0.913 | 0.918 | 1.000 | 0.830 | 0.964 |
| | **KOMAP** | **0.923** | **0.925** | **1.000** | **0.855** | **0.965** |
| | KOMAP separate [e] | 0.921 | 0.926 | 1.000 | 0.862 | 0.966 |
| AUPRC | PheCode290.11 | 0.835 | 0.836 | 0.792 | 0.825 | 0.841 |
| | PheCode290.1 | 0.659 | 0.663 | 0.933 | 0.536 | 0.729 |
| | PheCode290 | 0.621 | 0.642 | 0.460 | 0.502 | 0.682 |
| | C0002395 | 0.890 | 0.890 | 1.000 | 0.775 | 0.957 |
| | **KOMAP** | **0.904** | **0.903** | **1.000** | **0.868** | **0.926** |
| | KOMAP separate | 0.902 | 0.905 | 0.667 | 0.815 | 0.926 |
| TPR (at FPR≤ 0.1) | PheCode290.11 | 0.704 | 0.679 | 0.333 | 0.667 | 0.731 |
| | PheCode290.1 | 0.227 | 0.321 | 0.667 | 0.278 | 0.192 |
| | PheCode290 | 0.227 | 0.250 | 0.000 | 0.111 | 0.192 |
| | C0002395 | 0.659 | 0.786 | 1.000 | 0.500 | 0.769 |
| | **KOMAP** | **0.750** | **0.857** | **1.000** | **0.667** | **0.808** |
| | KOMAP separate | 0.750 | 0.857 | 1.000 | 0.667 | 0.808 |
| PPV (at FPR≤ 0.1) | PheCode290.11 | 0.886 | 0.864 | 1.000 | 0.857 | 0.905 |
| | PheCode290.1 | 0.667 | 0.750 | 1.000 | 0.714 | 0.714 |
| | PheCode290 | 0.667 | 0.778 | 1.000 | 0.667 | 0.714 |
| | C0002395 | 0.878 | 0.880 | 1.000 | 0.818 | 0.909 |
| | **KOMAP** | **0.868** | **0.889** | **1.000** | **0.857** | **0.913** |
| | KOMAP separate | 0.868 | 0.889 | 1.000 | 0.857 | 0.913 |
| NPV (at FPR≤ 0.1) | PheCode290.11 | 0.780 | 0.757 | 0.714 | 0.778 | 0.781 |
| | PheCode290.1 | 0.570 | 0.600 | 0.833 | 0.618 | 0.543 |
| | PheCode290 | 0.570 | 0.580 | 0.625 | 0.579 | 0.543 |
| | C0002395 | 0.754 | 0.824 | 1.000 | 0.700 | 0.806 |
| | **KOMAP** | **0.804** | **0.875** | **1.000** | **0.778** | **0.833** |
| | KOMAP separate | 0.804 | 0.875 | 1.000 | 0.778 | 0.833 |

a. We imputed missing race and ethnicity using multiple imputation based on age and gender.
b. NHW, non-Hispanic White individuals
c. Other, racial and ethnic minorities
d. Refer to S-Table 2 for explanation of PheCodes or Concept Unique Identifiers as benchmarks
e. “KOMAP separate” indicates that KOMAP was trained separately by race and ethnicity and by gender.

Other Abbreviations: *AUPRC*, area under the precision-recall curve; *AUROC,* area under the receiver operating characteristic curve; *FPR,* true positive rate; *NPV,* negative predictive value; *PPV*, positive predictive value; *TPR,* true positive rate.

**S-Table 10.** Comparative performance of AD diagnosis phenotyping algorithm (held at 90% specificity) against benchmarks at UPMC evaluated using the University of Pittsburgh ADRC registry labels.

| | **Method** | **All** | **Validation by Race and Ethnicity [a]** | | **Validation by Gender** | |
|---|---|---|---|---|---|---|
| | | | **NHW [b]** | **Other [c]** | **Men** | **Women** |
| AUROC | PheCode290.11 [d] | 0.794 | 0.788 | 0.836 | 0.784 | 0.803 |
| | PheCode290.1 [d] | 0.786 | 0.770 | 0.895 | 0.759 | 0.812 |
| | PheCode290 [d] | 0.780 | 0.763 | 0.896 | 0.753 | 0.805 |
| | C0002395 [d] | 0.719 | 0.704 | 0.814 | 0.717 | 0.725 |
| | **KOMAP** | 0.835 | 0.821 | 0.922 | 0.824 | 0.846 |
| | KOMAP separate [e] | 0.831 | 0.822 | 0.919 | 0.824 | 0.847 |
| AUPRC | PheCode290.11 | 0.829 | 0.823 | 0.870 | 0.771 | 0.864 |
| | PheCode290.1 | 0.765 | 0.750 | 0.880 | 0.705 | 0.816 |
| | PheCode290 | 0.758 | 0.743 | 0.879 | 0.701 | 0.807 |
| | C0002395 | 0.723 | 0.713 | 0.797 | 0.685 | 0.752 |
| | **KOMAP** | 0.833 | 0.824 | 0.906 | 0.784 | 0.867 |
| | KOMAP separate | 0.828 | 0.825 | 0.891 | 0.782 | 0.866 |
| TPR (at FPR≤ 0.1) | PheCode290.11 | 0.604 | 0.519 | 0.714 | 0.515 | 0.596 |
| | PheCode290.1 | 0.366 | 0.293 | 0.690 | 0.234 | 0.486 |
| | PheCode290 | 0.350 | 0.273 | 0.702 | 0.237 | 0.488 |
| | C0002395 | 0.348 | 0.307 | 0.607 | 0.320 | 0.339 |
| | **KOMAP** | 0.583 | 0.560 | 0.774 | 0.544 | 0.608 |
| | KOMAP separate | 0.575 | 0.560 | 0.762 | 0.544 | 0.613 |
| PPV (at FPR≤ 0.1) | PheCode290.11 | 0.865 | 0.852 | 0.846 | 0.824 | 0.876 |
| | PheCode290.1 | 0.795 | 0.764 | 0.842 | 0.680 | 0.852 |
| | PheCode290 | 0.788 | 0.752 | 0.844 | 0.683 | 0.853 |
| | C0002395 | 0.787 | 0.772 | 0.824 | 0.744 | 0.801 |
| | **KOMAP** | 0.861 | 0.861 | 0.856 | 0.832 | 0.878 |
| | KOMAP separate | 0.859 | 0.861 | 0.854 | 0.832 | 0.879 |
| NPV (at FPR≤ 0.1) | PheCode290.11 | 0.681 | 0.628 | 0.803 | 0.671 | 0.652 |
| | PheCode290.1 | 0.572 | 0.535 | 0.790 | 0.563 | 0.596 |
| | PheCode290 | 0.566 | 0.528 | 0.797 | 0.564 | 0.597 |
| | C0002395 | 0.565 | 0.540 | 0.748 | 0.592 | 0.534 |
| | **KOMAP** | 0.670 | 0.649 | 0.838 | 0.684 | 0.659 |
| | KOMAP separate | 0.666 | 0.649 | 0.831 | 0.684 | 0.662 |

a. We imputed missing race and ethnicity using multiple imputation based on age and gender.
b. NHW, non-Hispanic White individuals
c. Other, racial and ethnic minorities
d. Refer to S-Table 2 for explanation of PheCodes or Concept Unique Identifiers as benchmarks
e. "KOMAP separate" indicates that KOMAP was trained separately by race and ethnicity and by gender.

Other Abbreviations: *AUPRC*, area under the precision-recall curve; *AUROC,* area under the receiver operating characteristic curve; *FPR,* true positive rate; *NPV,* negative predictive value; *PPV*, positive predictive value; *TPR,* true positive rate.

**S-Table 11.** Comparison of AD diagnosis phenotyping algorithm performance (held at 90% specificity) evaluated using chart-reviewed labels at UPMC and MGB with and without imputation of race and ethnicity.

| | ***With* Imputation of Race and Ethnicity [b]** | | ***Without* Imputation of Race and Ethnicity [c]** | |
|---|---|---|---|---|
| | **UPMC** | **MGB** | **UPMC** | **MGB** |
| **AUROC** | 0.854 | 0.923 | 0.853 | 0.923 |
| **AUPRC** | 0.825 | 0.904 | 0.825 | 0.902 |
| **Sensitivity [a]** | 0.595 | 0.750 | 0.595 | 0.750 |
| **PPV [a]** | 0.839 | 0.868 | 0.840 | 0.868 |
| **NPV [a]** | 0.716 | 0.804 | 0.717 | 0.804 |

a. We calculated the sensitivity, PPV, and NPV at a specificity of 90%. (See Methods for rationale of holding specificity at 90%.)
b. AD cohort for all main analyses (see Main Table 1)
c. AD cohort for sensitivity analyses (see S-Table 9)

Other Abbreviations: *AUPRC*, area under the precision-recall curve; *AUROC,* area under the receiver operating characteristic curve; *NPV,* negative predictive value; *PPV*, positive predictive value.

**S-Table 12.** Performance of rule-based definitions of nursing home admission evaluated using chart-reviewed labels at UPMC and MGB.

| Site | Rule | Sensitivity TP/(TP + FN) | Specificity TN/(TN + FP) | PPV TP/(TP + FP) | NPV TN/(TN + FN) |
|---|---|---|---|---|---|
| **UPMC (n=100)** | ≥1 code | 0.493 | 0.968 | 0.971 | 0.462 |
| | ≥1 code or CUI | 0.913 | 0.903 | 0.955 | 0.824 |
| **MGB (n=100)** | ≥1 code | 0.709 | 1 | 1 | 0.738 |
| | ≥1 code or CUI | 0.945 | 0.911 | 0.929 | 0.932 |

Abbreviations: *FN:* false negative; *FP:* false positive; *NPV,* negative predictive value; *PPV*, positive predictive value; *TN:* true negative*; TP:* true positive.

**S-Table 13.** Cohort characteristics for sensitivity analyses (*without* imputation of race and ethnicity).

| | UPMC + MGB | UPMC | MGB | P-value |
|---|---|---|---|---|
| **Number of Patients, N (%)** | 24075 (100%) | 12938 (100%) | 11137 (100%) | |
| **Age at AD diagnosis, mean (SD)** | 79.55 (9.45) | 80.58 (9.01) | 78.35 (9.81) | **<.0001** |
| **Gender, N (%)** | | | | **<.0001** |
| Women | 14,782 (61%) | 8,248 (64%) | 6,534 (59%) | |
| Men | 9293 (39%) | 4690 (36%) | 4603 (41%) | |
| **Race and ethnicity [a], N (%)** | | | | **<.0001** |
| Non-Hispanic White | 21,702 (90%) | 11,872 (92%) | 9,830 (88%) | |
| American Indian | <20 (<0.1%) | <10 (<0.1%) | <20 (<0.1%) | |
| Asian | 280 (1.2%) | 59 (0.5%) | 221 (2.0%) | |
| Black | 1,442 (6.0%) | 964 (7.5%) | 478 (4.3%) | |
| Hawaiian or Pacific Islander | <10 (<0.1%) | <10 (<0.1%) | <10 (<0.1%) | |
| Hispanic or Latino | 389 (1.6%) | 36 (0.3%) | 353 (3.2%) | |
| Other | 243 (1.0%) | 0 (0%) | 243 (2.2%) | |
| **Baseline healthcare utilization (number of events) [b], mean (SD)** | 211.69 (247.23) | 188.70 (193.43) | 238.41 (295.55) | **<.0001** |
| **Baseline Elixhauser comorbidity index [b], mean (SD)** | | | | **<.0001** |
| <0 | 2,124 (8.8%) | 859 (7.7%) | 1,265 (9.8%) | |
| 0 | 5,028 (21%) | 2,845 (26%) | 2,183 (17%) | |
| 1-5 | 4,739 (20%) | 2,105 (19%) | 2,634 (20%) | |
| 6-13 | 7,154 (30%) | 3,328 (30%) | 3,826 (30%) | |
| 14+ | 5,030 (21%) | 2,000 (18%) | 3,030 (23%) | |
| **Baseline comorbidities (top 2 prevalent), N (%)** | | | | |
| Hypertension, uncontrolled | 15,456 (64%) | 9,313 (72%) | 6,143 (55%) | **<.0001** |
| Cardiac arrhythmia | 7,853 (33%) | 4,565 (35%) | 3,288 (30%) | **<.0001** |
| **Survival, N (%)** | | | | **<.0001** |
| Alive | 12,258 (51%) | 5,106 (39%) | 7,152 (64%) | |
| Death | 11,817 (49%) | 7,832 (61%) | 3,985 (36%) | |
| **Time to death, median months (1st and 3rd quantiles)** | 32 (15, 56) | 33 (16, 54) | 31 (14, 59) | 0.449 |
| **Nursing Home Admission, N (%)** | | | | **<.0001** |
| Admitted | 12,545 (52%) | 8,555 (66%) | 3,990 (36%) | |
| Not admitted | 11,530 (48%) | 4,383 (34%) | 7,147 (64%) | |
| **Time to nursing home admission, median months (1st and 3rd quantiles)** | 21 (7, 42) | 21 (7, 41) | 22 (8, 45) | **0.002** |
| **AD-related medication [c], N (%)** | | | | |
| Not prescribed | 5,566 (23%) | 2,469 (19%) | 3,097 (28%) | **<.0001 [d]** |
| Prescribed | 18,509 (77%) | 10,469 (81%) | 8,040 (72%) | |
| Before AD diagnosis | 9,591 (52%) | 5,766 (55%) | 3,825 (48%) | **<.0001 [e]** |
| On or after AD diagnosis | 8,918 (48%) | 4,703 (45%) | 4,215 (52%) | |
| **Follow-up duration [f], median months (1st and 3rd quantiles)** | 76 (42, 116) | 83 (56, 118) | 64 (29, 112) | **<.0001** |

a. We *did not* impute missing race and ethnicity and excluded patients without race and ethnicity.
b. We calculated the baseline healthcare utilization and comorbidity burden using the Elixhauser index (with van Walraven weighting) in the 24 months prior to AD diagnosis.

c. The complete list of AD-related medications is available in S-Table 3.
d. We used a two-sample t-test to compare the difference between the number of patients prescribed and not prescribed AD-related medication.
e. We used a two-sample t-test to compare the difference between the number of patients prescribed AD-related medication before vs after AD diagnosis.
f. We estimated the median follow-up duration using the reverse Kaplan-Meier survival analysis.

**S-Table 14.** Adjusted hazard ratios in the fixed-effects meta-analysis of healthcare system-specific covariate-adjusted Cox proportional hazard models of **nursing home admission** *with* and *without* imputation of race and ethnicity.

| **Baseline Features** | **Meta-analysis [a]** | | **Meta-analysis [b]** | |
|---|---|---|---|---|
| | **Hazard Ratio (95% CI)** | **P-value** | **Hazard Ratio (95% CI)** | **P-value** |
| psychoses | 1.295 (1.226, 1.369) | **<.0001** | 1.330 (1.252, 1.411) | **<.0001** |
| hypertension, uncomplicated | 1.184 (1.135, 1.234) | **<.0001** | 1.186 (1.134, 1.241) | **<.0001** |
| diabetes, uncontrolled | 1.137 (1.085, 1.191) | **<.0001** | 1.144 (1.090, 1.202) | **<.0001** |
| chronic pulmonary disease | 1.112 (1.065, 1.161) | **<.0001** | 1.101 (1.052, 1.153) | **<.0001** |
| fluid and electrolyte disorders | 1.105 (1.057, 1.154) | **<.0001** | 1.118 (1.067, 1.171) | **<.0001** |
| depression | 1.085 (1.045, 1.127) | **<.0001** | 1.093 (1.050, 1.138) | **<.0001** |
| healthcare utilization: log (util + 1) | 1.051 (1.029, 1.074) | **<.0001** | 1.028 (1.004, 1.052) | **0.023** |
| age at AD diagnosis | 1.021 (1.019, 1.023) | **<.0001** | 1.023 (1.020, 1.025) | **<.0001** |
| other neurological disorders | 0.929 (0.892, 0.967) | **<.0001** | 0.942 (0.902, 0.983) | **0.007** |
| hypertension, complicated | 1.102 (1.040, 1.169) | **0.001** | 1.100 (1.031, 1.173) | **0.004** |
| weight loss | 1.072 (1.018, 1.129) | **0.008** | 1.076 (1.018, 1.137) | **0.009** |
| gender: women | 1.061 (1.024, 1.100) | **0.001** | 1.065 (1.025, 1.107) | **0.001** |
| paralysis | 1.162 (1.011, 1.336) | **0.035** | 1.156 (0.993, 1.346) | 0.062 |
| rheumatoid arthritis | 1.088 (1.015, 1.166) | **0.017** | 1.083 (1.006, 1.166) | **0.033** |
| renal failure | 1.064 (1.003, 1.128) | **0.040** | 1.062 (0.997, 1.131) | 0.060 |
| peripheral vascular disorder | 1.062 (1.012, 1.113) | **0.014** | 1.050 (0.998, 1.105) | 0.062 |
| drug abuse | 1.136 (0.983, 1.313) | 0.084 | 1.184 (1.018, 1.376) | **0.029** |
| coagulopathy | 1.078 (0.997, 1.165) | **0.058** | 1.085 (0.999, 1.178) | 0.052 |
| alcohol abuse | 1.063 (0.953, 1.185) | 0.274 | 1.055 (0.938, 1.186) | 0.374 |
| peptic ulcer disease | 1.044 (0.941, 1.158) | 0.417 | 1.030 (0.923, 1.149) | 0.598 |
| congestive heart failure | 1.042 (0.987, 1.100) | 0.139 | 1.045 (0.986, 1.108) | 0.134 |
| deficiency anemia | 1.034 (0.976, 1.096) | 0.251 | 1.025 (0.964, 1.089) | 0.431 |
| cardiac arrhythmias | 1.032 (0.992, 1.074) | 0.120 | 1.044 (1.001, 1.090) | **0.045** |
| valvular disease | 1.031 (0.983, 1.082) | 0.206 | 1.037 (0.986, 1.092) | 0.158 |
| obesity | 1.019 (0.952, 1.091) | 0.584 | 1.019 (0.949, 1.095) | 0.598 |
| hypothyroidism | 1.017 (0.976, 1.060) | 0.415 | 1.023 (0.980, 1.069) | 0.302 |
| liver disease | 1.008 (0.929, 1.093) | 0.856 | 0.994 (0.913, 1.082) | 0.887 |
| race/ethnicity: non-Hispanic white | 1.006 (0.952, 1.063) | 0.831 | 1.009 (0.952, 1.069) | 0.767 |
| metastatic cancer | 0.999 (0.880, 1.134) | 0.987 | 1.027 (0.896, 1.178) | 0.700 |
| solid tumor, without metastasis | 0.994 (0.946, 1.044) | 0.801 | 0.986 (0.935, 1.040) | 0.608 |
| lymphoma | 0.983 (0.846, 1.142) | 0.823 | 0.980 (0.835, 1.151) | 0.808 |
| diabetes, controlled | 0.969 (0.908, 1.035) | 0.349 | 0.960 (0.895, 1.029) | 0.250 |
| pulmonary circulation disorders | 0.939 (0.864, 1.020) | 0.137 | 0.938 (0.860, 1.022) | 0.145 |
| blood loss anemia | 0.927 (0.806, 1.065) | 0.285 | 0.971 (0.842, 1.120) | 0.686 |
| AIDS/HIV | 0.839 (0.407, 1.731) | 0.634 | 1.000 (0.485, 2.064) | 0.999 |

a. We imputed missing race and ethnicity using multiple imputation based on age and gender.
b. We excluded the patients with missing race and ethnicity information.

Note: The fixed-effects meta-analysis model was adjusted for pooled covariates across both healthcare systems from inverse-probability weighting. Covariates included demographics (*e.g.*, age at AD diagnosis, gender, race, ethnicity) and baseline clinical profiles (*e.g.,* healthcare utilization, pre-existing comorbidity burden) in the 24 months preceding the index date.

**S-Table 15.** Adjusted hazard ratios in the fixed-effects meta-analysis of healthcare system-specific covariate-adjusted Cox proportional hazard models of **death** *with* and *without* imputation of race and ethnicity.

| **Baseline Features** | **Meta-analysis [a]** | | **Meta-analysis [b]** | |
|---|---|---|---|---|
| | **Hazard Ratio (95% CI)** | **P-value** | **Hazard Ratio (95% CI)** | **P-value** |
| metastatic cancer | 1.881 (1.587, 2.231) | **<.0001** | 1.774 (1.436, 2.191) | **<.0001** |
| race/ethnicity: non-Hispanic white | 1.376 (1.245, 1.521) | **<.0001** | 1.458 (1.283, 1.657) | **<.0001** |
| congestive heart failure | 1.373 (1.257, 1.500) | **<.0001** | 1.333 (1.197, 1.484) | **<.0001** |
| psychoses | 1.370 (1.250, 1.500) | **<.0001** | 1.240 (1.099, 1.399) | **<.0001** |
| peripheral vascular disorder | 1.206 (1.112, 1.309) | **<.0001** | 1.168 (1.059, 1.289) | **0.002** |
| diabetes, uncontrolled | 1.197 (1.103, 1.299) | **<.0001** | 1.153 (1.044, 1.273) | **0.005** |
| weight loss | 1.188 (1.090, 1.295) | **<.0001** | 1.192 (1.074, 1.323) | **0.001** |
| age at AD diagnosis | 1.018 (1.015, 1.022) | **<.0001** | 1.018 (1.014, 1.023) | **<.0001** |
| gender: women | 0.856 (0.811, 0.904) | **<.0001** | 0.823 (0.769, 0.882) | **<.0001** |
| healthcare utilization: log (util + 1) | 0.726 (0.706, 0.747) | **<.0001** | 0.792 (0.762, 0.822) | **<.0001** |
| deficiency anemia | 1.207 (1.091, 1.336) | **<.0001** | 1.192 (1.057, 1.344) | **0.004** |
| solid tumor, without metastasis | 1.154 (1.068, 1.248) | **<.0001** | 1.146 (1.042, 1.261) | **0.005** |
| obesity | 0.780 (0.680, 0.893) | **<.0001** | 0.866 (0.742, 1.010) | 0.067 |
| lymphoma | 1.423 (1.138, 1.779) | **0.002** | 1.523 (1.187, 1.955) | **0.001** |
| coagulopathy | 1.210 (1.062, 1.378) | **0.004** | 1.153 (0.983, 1.351) | 0.079 |
| chronic pulmonary disease | 1.119 (1.038, 1.206) | **0.003** | 1.087 (0.993, 1.190) | 0.072 |
| cardiac arrhythmias | 1.111 (1.040, 1.187) | **0.002** | 1.034 (0.954, 1.122) | 0.415 |
| depression | 0.914 (0.854, 0.978) | **0.009** | 0.867 (0.799, 0.942) | **0.001** |
| diabetes, controlled | 0.831 (0.736, 0.938) | **0.003** | 0.883 (0.766, 1.017) | 0.084 |
| fluid and electrolyte disorders | 1.098 (1.015, 1.188) | **0.020** | 1.037 (0.945, 1.139) | 0.440 |
| hypertension, uncomplicated | 1.078 (1.011, 1.149) | **0.023** | 1.049 (0.969, 1.136) | 0.234 |
| renal failure | 0.897 (0.810, 0.993) | **0.037** | 0.989 (0.880, 1.112) | 0.859 |
| AIDS/HIV | 1.425 (0.452, 4.493) | 0.545 | 1.622 (0.508, 5.182) | 0.414 |
| peptic ulcer disease | 1.117 (0.909, 1.373) | 0.293 | 1.105 (0.865, 1.411) | 0.424 |
| pulmonary circulation disorders | 1.084 (0.930, 1.263) | 0.305 | 1.134 (0.953, 1.350) | 0.157 |
| rheumatoid arthritis | 1.073 (0.947, 1.216) | 0.269 | 0.993 (0.850, 1.160) | 0.927 |
| valvular disease | 1.038 (0.954, 1.129) | 0.388 | 0.999 (0.902, 1.106) | 0.977 |
| paralysis | 1.025 (0.808, 1.301) | 0.839 | 1.013 (0.750, 1.369) | 0.933 |
| blood loss anemia | 1.025 (0.796, 1.319) | 0.849 | 1.107 (0.837, 1.465) | 0.476 |
| liver disease | 1.015 (0.870, 1.183) | 0.852 | 0.946 (0.787, 1.137) | 0.554 |
| other neurological disorders | 1.011 (0.949, 1.078) | 0.728 | 0.898 (0.827, 0.975) | **0.010** |
| hypothyroidism | 0.994 (0.923, 1.070) | 0.876 | 0.976 (0.893, 1.066) | 0.590 |
| hypertension, complicated | 0.966 (0.878, 1.063) | 0.476 | 0.934 (0.829, 1.052) | 0.261 |
| alcohol abuse | 0.939 (0.778, 1.133) | 0.512 | 0.941 (0.741, 1.195) | 0.615 |
| drug abuse | 0.763 (0.557, 1.046) | 0.093 | 0.601 (0.391, 0.923) | **0.020** |

a. We imputed missing race and ethnicity using multiple imputation based on age and gender.
b. We excluded the patients with missing race and ethnicity information.

Note: The fixed-effects meta-analysis model was adjusted for pooled covariates across both healthcare systems from inverse-probability weighting. Covariates included demographics (*e.g.*, age at AD diagnosis, gender, race, ethnicity) and baseline clinical profiles (*e.g.,* healthcare utilization, pre-existing comorbidity burden) in the 24 months preceding the index date.

### SUPPLEMENTAL FIGURES

**S-Figure 1.** Inclusion criteria at UPMC and MGB. (A) With imputation of missing race and ethnicity. (B) Without imputation of missing race and ethnicity.

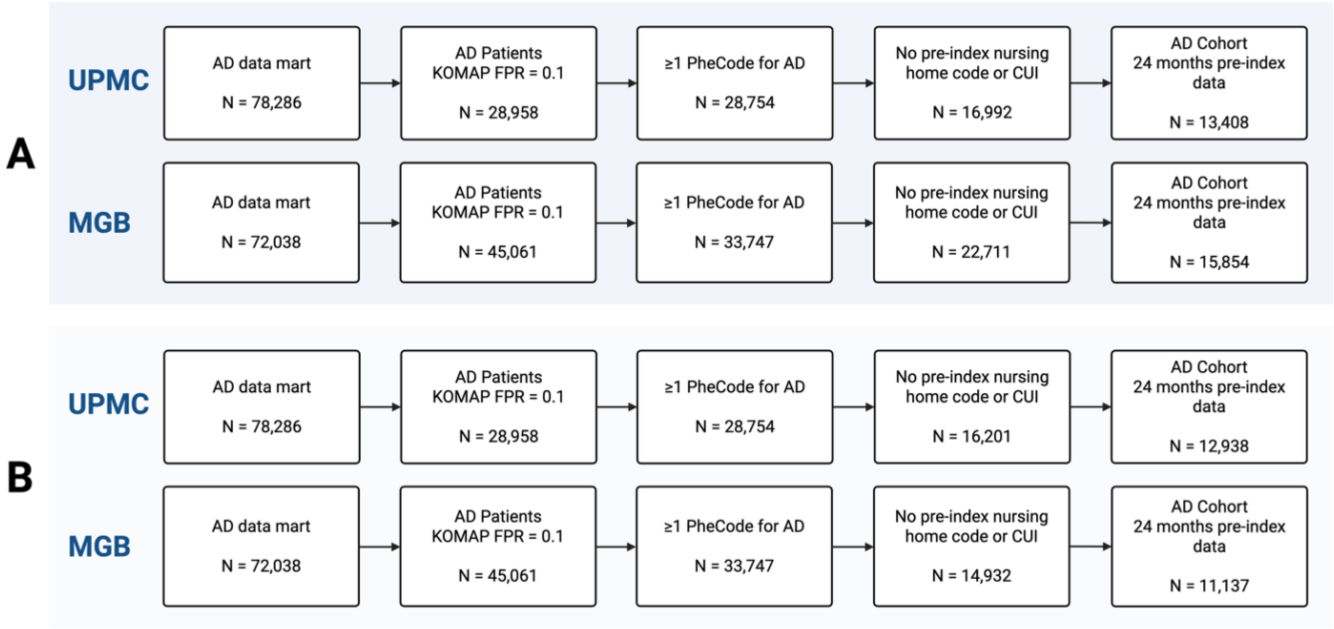

**S-Figure 2.** Comorbidity burden by race and ethnicity and by gender. (A) Combined main AD cohort *with* imputation of race and ethnicity (B) Combined sensitivity analysis AD cohort *without* imputation of race and ethnicity. Variables are ordered by the percentage of patients with the comorbidity then by p-value significance threshold (*p*<.05*, *p*<.01**, *p*<.001***, *p*<.0001****).

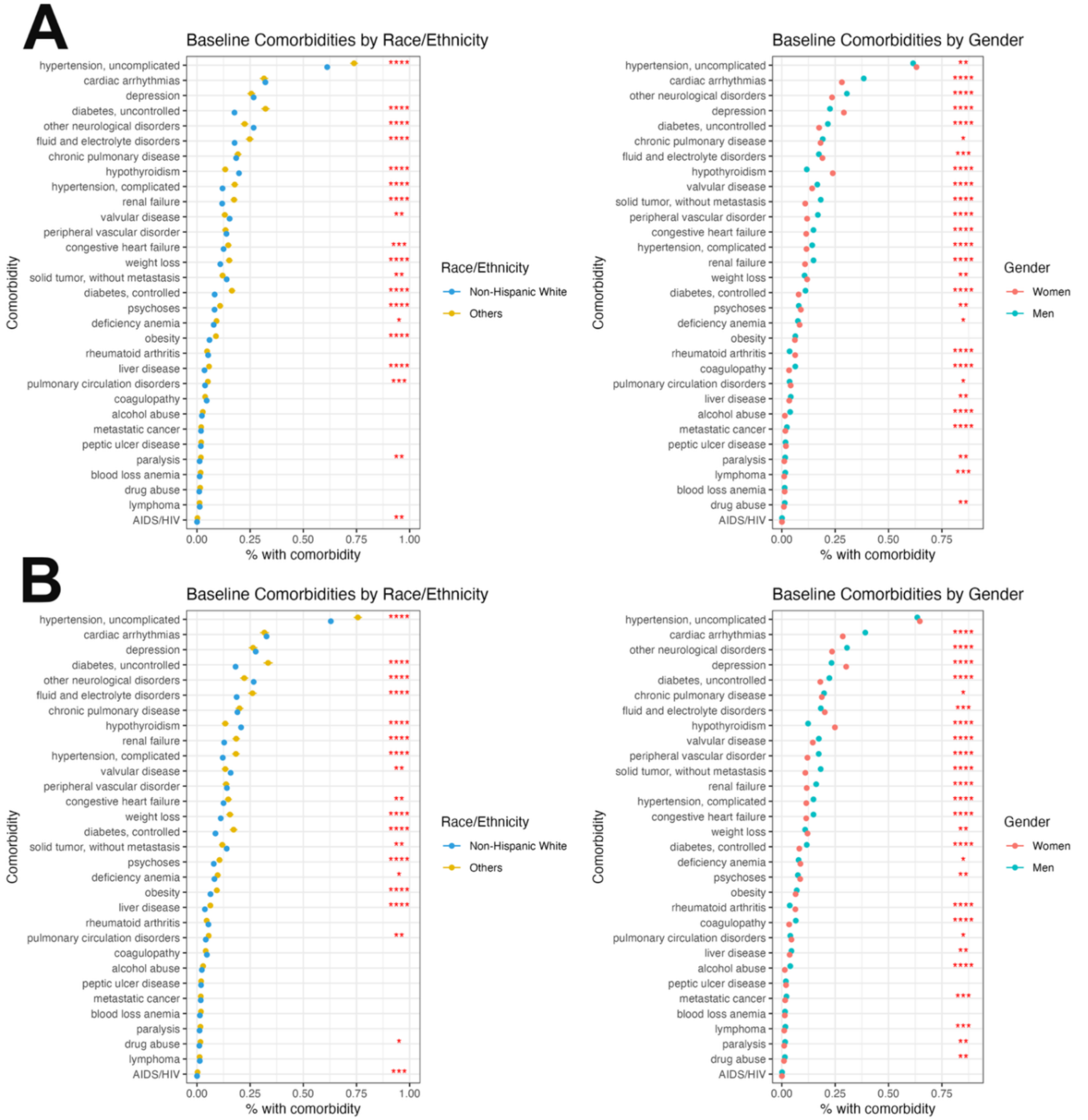


P-value significance threshold: *p*<.05*, *p*<.01**, *p*< .001***, and *p*<.0001****

**S-Figure 3.** Fixed-effects meta-analysis of healthcare system-specific covariate-adjusted Cox proportional hazard models to estimate time to nursing home admission and death by demographic groups (using van Walraven weighted Elixhauser score). The fixed-effects meta-analysis model was adjusted for pooled covariates across both healthcare systems from inverse-probability weighting. Covariates included demographics (*e.g.*, age at AD diagnosis, gender, race, ethnicity) and baseline clinical profiles (*e.g.,* healthcare utilization, pre-existing comorbidity burden) in the 24 months preceding the index date. We accounted for pre-existing comorbidity burden using the van Walraven weighted Elixhauser score. (A-B) Nursing Home Admission. (C-D) Death. Reference group for race and ethnicity: non-Hispanic White. Reference group for gender: women.

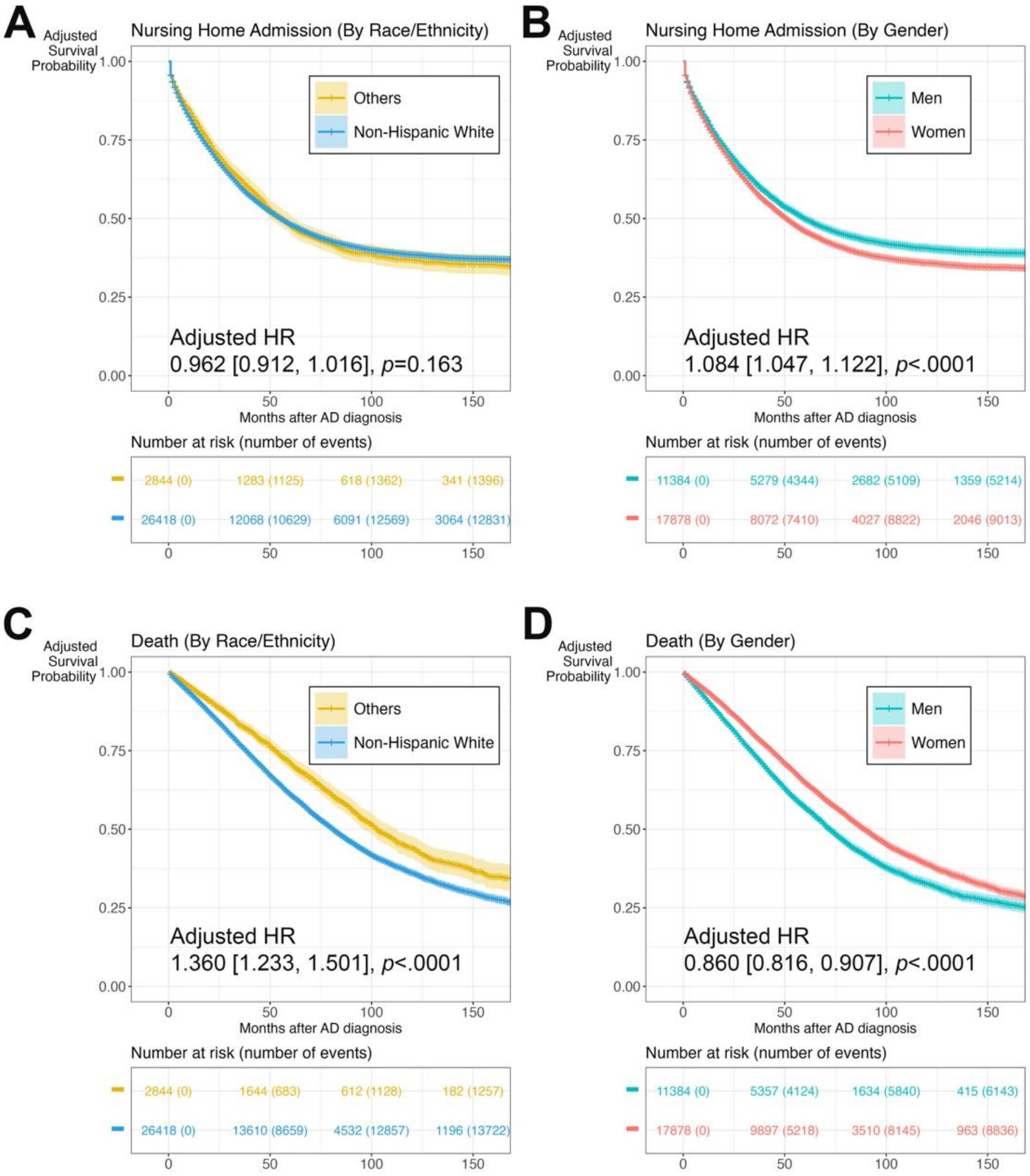

**S-Figure 4.** Adjusted hazard ratios of variables in the fixed-effects meta-analysis of healthcare system-specific covariate-adjusted Cox proportional hazard models of nursing home admission and death (using van Walraven weighted Elixhauser score). The fixed-effects meta-analysis model was adjusted for pooled covariates across both healthcare systems from inverse-probability weighting. Covariates included demographics (*e.g.*, age at AD diagnosis, gender, race, ethnicity) and baseline clinical profiles (*e.g.,* healthcare utilization, pre-existing comorbidity burden) in the 24 months preceding the index date. We accounted for pre-existing comorbidity burden using the van Walraven weighted Elixhauser score. Variables are ordered first by p-value significance threshold ($p$<.05*, $p$<.01**, $p$<.001***, $p$<.0001****) and then effect size. (A) Nursing Home Admission. (B) Death. Reference group for race and ethnicity: non-Hispanic White. Reference group for gender: women.

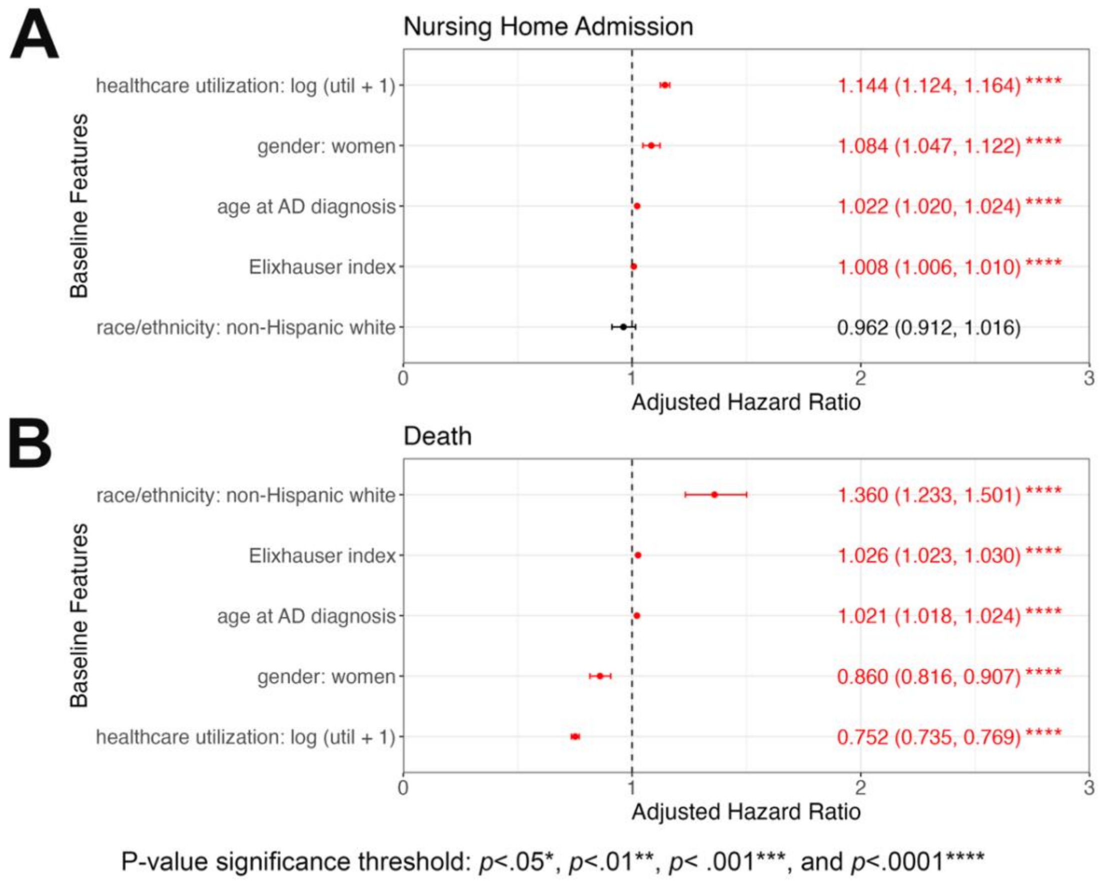

**S-Figure 5.** Fixed-effects meta-analysis of healthcare system-specific covariate-adjusted Cox proportional hazard models to estimate time to nursing home admission and death by demographic groups (*without* imputation of race and ethnicity). We excluded patients with missing race and ethnicity information. The fixed-effects meta-analysis model was adjusted for pooled covariates across both healthcare systems from inverse-probability weighting. Covariates included demographics (*e.g.*, age at AD diagnosis, gender, race, ethnicity) and baseline clinical profiles (*e.g.,* healthcare utilization, pre-existing comorbidity burden) in the 24 months preceding the index date. (A-B) Nursing Home Admission. (C-D) Death. Reference group for race and ethnicity: non-Hispanic White. Reference group for gender: women.

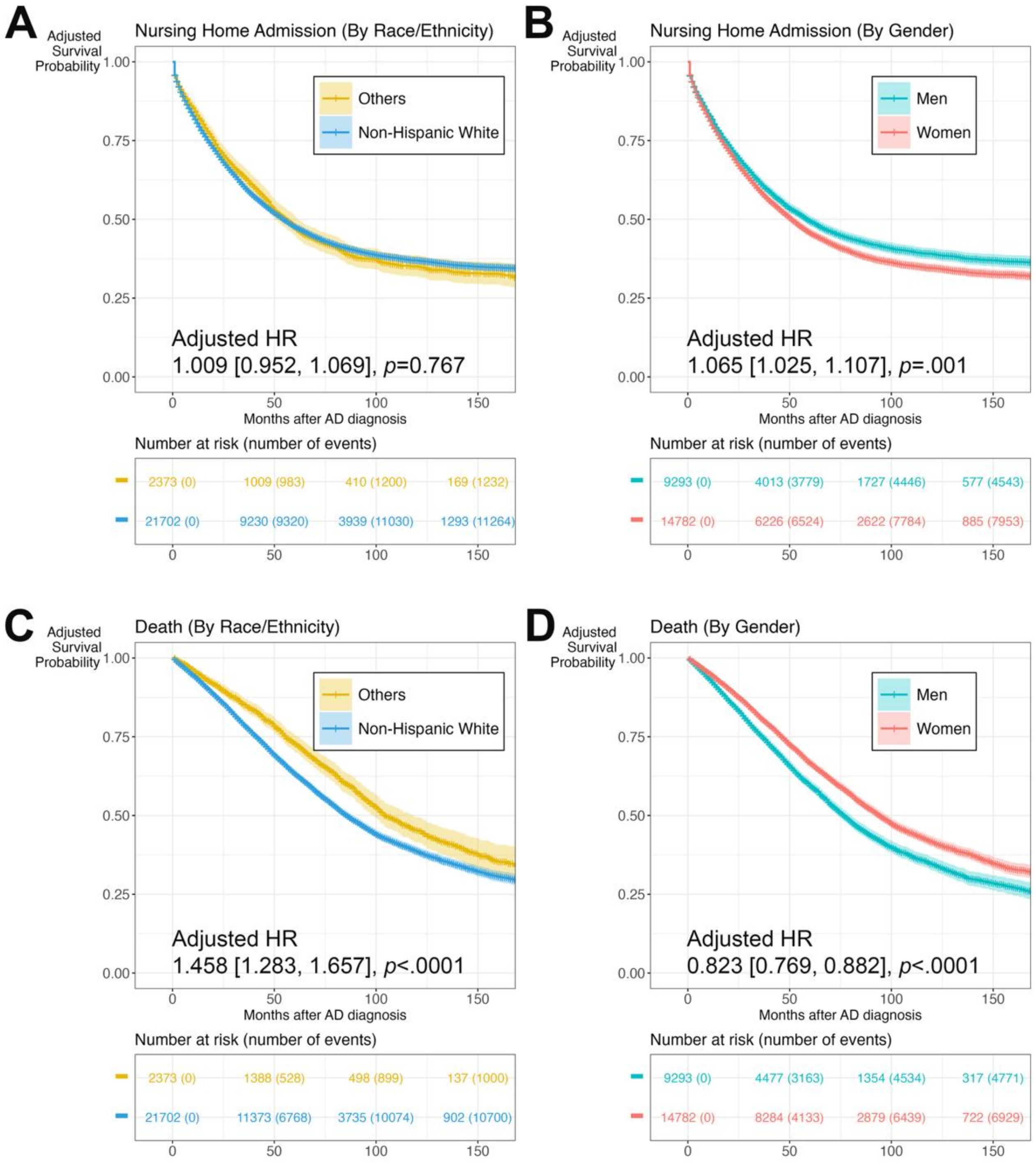

**S-Figure 6.** Adjusted hazard ratios of variables in the fixed-effects meta-analysis of healthcare system-specific covariate-adjusted Cox proportional hazard models of nursing home admission and death (*without* imputation of race and ethnicity). We excluded patients with missing race and ethnicity information. The fixed-effects meta-analysis model was adjusted for pooled covariates across both healthcare systems from inverse-probability weighting. Covariates included demographics (*e.g.*, age at AD diagnosis, gender, race, ethnicity) and baseline clinical profiles (*e.g.,* healthcare utilization, pre-existing comorbidity burden) in the 24 months preceding the index date. We accounted for pre-existing comorbidity burden using the van Walraven weighted Elixhauser score. Variables are ordered first by p-value significance threshold ($p$<.05*, $p$<.01**, $p$<.001***, $p$<.0001****) and then effect size. (A) Nursing Home Admission. (B) Death.

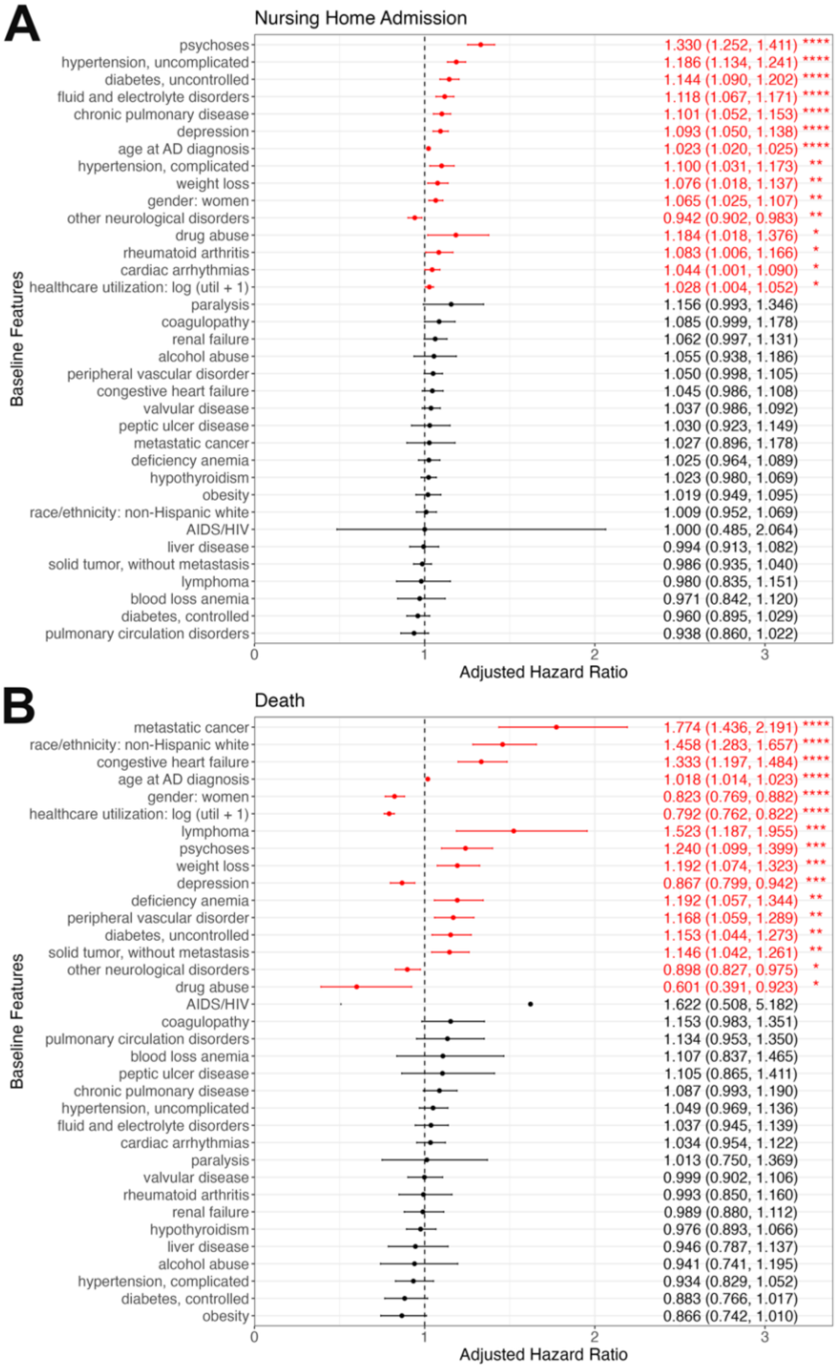


P-value significance threshold: $p$<.05*, $p$<.01**, $p$< .001***, and $p$<.0001****
